# Standardized transcriptome analysis improves rare disease diagnosis in the pan-European Solve-RD consortium

**DOI:** 10.64898/2026.02.10.26345954

**Authors:** Vicente A. Yépez, Rebeka Luknárová, Danique Beijer, Berta Estévez-Arias, Davide Mei, Heba Morsy, Juliane S. Mueller, Kiran Polavarapu, German Demidov, Cenna Doornbos, Kornelia Ellwanger, Leon Kraß, Steven Laurie, Leslie Matalonga, Ibrahim M. Abdelrazek, Galuh Astuti, Francesca Bisulli, Felix Brechtmann, Marc Dabad, Anne-Sophie Denommé-Pichon, Minas Drakos, Zakaria Eddafir, Glòria Garrabou, Renzo Guerrini, Mridul Johari, Josua Kegele, Ozge Aksel Kilicarslan, Heike Koelbel, Ingrid H.M. Kolen, Laura Licchetta, Hanns Lochmüller, Kars Maassen, William Macken, Christian Mertes, José César Milisenda, Raffaella Minardi, Barbara Mostacci, Kornelia Neveling, Machteld M. Oud, Joohyun Park, Aurora Pujol, Andreas Roos, Lydia Sagath, Bart van der Sanden, Marco Savarese, Alba Segarra-Casas, Noor Smal, Marloes Steehouwer, Wouter Steyaert, Rachel Thompson, Mirja Thomsen, Gerben van der Vries, Carlo Wilke, Ioannis Zaganas, Sergi Beltran, Alexander Hoischen, Katja Lohmann, Daniel Natera-de Benito, Rebecca Schüle, Matthis Synofzik, Lisenka Vissers, Ana Töpf, Holm Graessner, Antonio Vitobello, Peter-Bram ’t Hoen, Solve-RD Consortium, Anna Esteve-Codina, Julien Gagneur

**Affiliations:** School of Computation, Information and Technology, Technical University of Munich, Garching, Germany; Division Translational Genomics of Neurodegenerative Diseases, Hertie-Institute for Clinical Brain Research and Center of Neurology, University of Tübingen, Tübingen, Germany; Neuromuscular Unit, Hospital Sant Joan de Déu, Barcelona, Spain; Applied Research in Neuromuscular Diseases, Institut de Recerca Sant Joan de Déu, Barcelona, Spain; Department of Neuroscience and Human Genetics, Meyer Children’s Hospital IRCCS, Florence, Italy; Department of Neuromuscular Diseases, UCL Queen Square Institute of Neurology, London, UK; Department of Human Genetics, Medical Research Institute, Alexandria University, Egypt; Dubowitz Neuromuscular Centre, UCL Great Ormond Street Hospital, London, UK; Children’s Hospital of Eastern Ontario Research Institute, Ottawa, Canada; Institute of Medical Genetics and Applied Genomics, University of Tübingen, Tübingen, Germany; Department of Medical BioSciences, Radboud University Medical Center, Nijmegen, The Netherlands; Institute of Human Genetics, School of Medicine, Technical University of Munich, Munich, Germany; Computational Health Center, Helmholtz Center Munich, Neuherberg, Germany; Centro Nacional de Análisis Genómico (CNAG), Barcelona, Spain; Universitat de Barcelona (UB), Barcelona, Spain; Department of Human Genetics, Research Institute for Medical Innovation, Radboud University Medical Center, Nijmegen, the Netherlands; IRCCS Istituto delle Scienze Neurologiche di Bologna, Full Member of European Reference Network EpiCARE, Bologna, Italy; Department of Biomedical and Neuromotor Sciences, University of Bologna, Bologna, Italy; Medical genomics, FHU TRANSLAD, Dijon University Hospital, Dijon, France; Inserm - Burgundy Europe University, unit 1231 GAD team, Dijon, France; Medical School, University of Crete; Neurology Department, University Hospital of Heraklion, Crete; Applied and Translational Neurogenomics Group, VIB Center for Molecular Neurology, VIB, Antwerp, Belgium; Applied Translational Neurogenomics Group, Department of Biomedical Sciences, University of Antwerp, Antwerp, Belgium; Inherited Metabolic Disorders and Muscular Diseases Research Group, Cellex-IDIBAPS and Faculty of Medicine and Health Sciences, University of Barcelona, Barcelona, Spain; Department of Internal Medicine, Hospital Clinic of Barcelona, Barcelona, Spain; Centre for Biomedical Research on Rare Diseases (CIBERER), Instituto de Salud Carlos III, Madrid, Spain; Neuroscience Department, University of Florence, Florence, Italy; Folkhälsan Research Centre and Medicum, University of Helsinki, Helsinki, Finland; Harry Perkins Institute of Medical Research, Centre for Medical Research, University of Western Australia, Nedlands, WA, Australia; Department of Cellular and Molecular Medicine, University of Ottawa, ON, Canada; University Medicine Essen, Department of Pediatric Neurology, Centre for Neuromuscular Disorders, University Duisburg-Essen, Essen, Germany; Division of Neurodegenerative Diseases and Movement Disorders, Department of Neurology, Heidelberg University Hospital and Faculty of Medicine, Heidelberg, Germany; Division of Neurology, Department of Medicine, The Ottawa Hospital, Ottawa, Ontario, Canada; Brain and Mind Research Institute, Children’s Hospital of Eastern Ontario Research Institute, University of Ottawa, Ottawa, Canada; Department of Genetics, University Medical Center Groningen, University of Groningen, Groningen, The Netherlands; Neurometabolic Diseases Laboratory, Bellvitge Biomedical Research Institute (IDIBELL), L’Hospitalet de Llobregat, Barcelona, Catalonia, Spain; Catalan Institution of Research and Advanced Studies (ICREA), Barcelona, Catalonia, Spain; Department of Neurology with Heimer Institute for Muscle Research, University Hospital Bergmannsheil, Bochum, Germany; John Walton Muscular Dystrophy Research Centre, Translational and Clinical Research Institute, Newcastle University and Newcastle Hospitals NHS Foundation Trust, Newcastle upon Tyne, UK; Genetics Department, Institut de Recerca Sant Pau (IR SANT PAU), Hospital de la Santa Creu i Sant Pau, Barcelona, Spain; Radboud Institute for Molecular Life Sciences, Nijmegen, The Netherlands; Institute of Neurogenetics, University of Lübeck, Lübeck, Germany; Department of Genetics, Genomics Coordination Center, University Medical Center Groningen, University of Groningen, Groningen, The Netherlands; German Center for Neurodegenerative Diseases (DZNE), Tübingen, Germany; Departament de Genètica, Microbiologia i Estadística, Facultat de Biologia, Universitat de Barcelona (UB), Barcelona, Spain; Department of Internal Medicine and Radboud Center for Infectious Diseases (RCI), Radboud University Medical Center, Nijmegen, the Netherlands; Centre for Rare Diseases, University of Tübingen, Tübingen, Germany

## Abstract

RNA sequencing (RNA-seq) provides a powerful complement to DNA sequencing for uncovering pathogenic defects affecting gene expression and splicing in individuals with genetically undiagnosed rare disorders. However, as large rare disease consortia adopt RNA-seq, challenges arise due to cohort heterogeneity, variability in tissues and sample sizes, and differences in interpretation practices.

Here, we present a harmonized analytical and interpretation framework developed by the pan-European Solve-RD consortium to address these challenges. We analyzed 521 RNA-seq samples from whole blood, fibroblasts, muscle and peripheral blood mononuclear cells collected across more than 30 clinics and five European Reference Networks. Aberrant expression and splicing events were identified using OUTRIDER and FRASER 2.0 and analysed through a standardized four-level scoring framework that encompassed RNA-seq outlier reliability, phenotype relevance, variant mechanism, and segregation evidence, captured in structured reports for interpretation. Regular meetings, and collaborative “Solvathon” workshops were used to evaluate variant pathogenicity.

This effort resulted in molecular diagnoses for 19 families out of 248 (7.7%) for whom DNA analyses had been inconclusive. Furthermore, three cases diagnosed using DNA analyses were confirmed, and 49 candidate events and five novel candidate disease genes were identified in the remaining families. Our results demonstrate the feasibility and impact of large-scale, standardized RNA-seq analysis in a transnational research setting. This framework provides a model for other international initiatives such as the Undiagnosed Diseases Network and ERDERA, paving the way for broader clinical implementation of transcriptome-based rare disease diagnostics.

## Introduction

Rare diseases, which collectively affect millions of individuals worldwide, remain genetically unexplained in approximately half of affected individuals despite comprehensive genomic testing (The 100,000 Genomes Project Pilot Investigators, 2021; Boycott et al., 2022; Stark et al., 2023; Lindstrand et al., 2025). Short-read DNA sequencing technologies such as exome and genome sequencing have paved the way as first-tier diagnostic tools by identifying variants that may underlie rare conditions (Yang et al., 2013; Wojcik et al., 2024). As our knowledge on gene and variant level continues to improve, periodic reanalysis of these data is very valuable, as previously demonstrated by Solve-RD (Laurie et al., 2025). In addition, recent advances, such as long-read genome sequencing and optical genome mapping, have substantially improved the detection of structural and copy-number variants, repeat expansions, and other complex genomic alterations (Miller et al., 2021; Liu et al., 2024; Höps et al., 2025; Steyaert et al., 2025). However, the interpretation of these structural variants, as well as single-nucleotide variants (SNVs) and short indels in non-coding and regulatory regions, remains a major bottleneck in the diagnostic workflow (Cohen et al., 2021; Martin-Geary et al., 2025).

RNA sequencing (RNA-seq) has emerged as a powerful complement to DNA-based analyses by providing a functional readout of gene activity (Cummings et al., 2017; Kremer et al., 2017; Montgomery et al., 2022; Zhao et al., 2025). It can uncover the molecular consequences of genetic variants by detecting aberrant gene expression and splicing. In terms of analysis workflow, RNA-seq can be used in two main ways: 1) DNA-first approach, for which RNA-seq serves as a functional assay to validate or interpret variants of uncertain significance (VUS) from DNA sequencing; and 2) RNA-first approach, for which new candidate pathogenic events are discovered from a transcriptome-wide analysis pointing to genes that were missed, not captured, or not prioritized by DNA analysis alone (Marwaha et al., 2022; Tesi et al., 2023; Kernohan and Boycott, 2024).

Early studies combined RNA-seq with SNVs and small indels called from exome or genome sequencing in a broad range of rare genetic disorders and increased diagnostic yields by up to 36% using clinically-accessible tissues such as skeletal muscle, fibroblasts, or blood (Cummings et al., 2017; Kremer et al., 2017; Frésard et al., 2019; Gonorazky et al., 2019; Murdock et al., 2021). More recently, studies have expanded the range of usable tissues, including transdifferentiated cells (Li et al., 2024), amniotic fluid cells (Lee et al., 2022), and dried blood spots (Bertoli-Avella et al., 2025), expanding the applicability of transcriptome analysis. Recent work has also shown that integrating expression and splicing insights with structural variants, such as deletions, duplications and translocations, can improve their interpretation, increase the diagnostic yield and mechanistic insight (Vialle et al., 2022; Lunke et al., 2023; Xiao et al., 2024; Jensen et al., 2025). Altogether, the added value of RNA-seq has been demonstrated by uplifts in diagnostic rates of 3 to 36% across various centers, tissues and diseases, including mitochondrial, neuromuscular, skin-related, inborn errors of metabolism, and epilepsies and other neurological disorders (Youssefian et al., 2021; Hong et al., 2022; Yépez et al., 2022; Deshwar et al., 2023; Jaramillo Oquendo et al., 2024; Marchant et al., 2024; Riquin et al., 2024; Arriaga et al., 2025; De Cock et al., 2025; Luo et al., 2025; Segarra-Casas et al., 2025; Stark et al., 2025; Saparov et al., 2026). Nonetheless, scaling such analyses across heterogeneous disease cohorts and tissues remains challenging, particularly for large multi-center initiatives, due to variability in data and RNA quality, tissue specificity, and interpretation practices.

The Solve-Rare Diseases Consortium (Solve-RD), a pan-European initiative, was established to increase diagnostic rates in rare diseases by reanalyzing previously generated short read DNA sequencing data (Laurie et al., 2025) and analyzing newly produced data, including long-read genome sequencing (Steyaert et al., 2025), deep exome sequencing (Sommer et al., 2026), optical genome mapping, RNA-seq, and epigenetics data. The consortium is composed of i) sample submitters and data interpreters (organized into Data Interpretation Task Forces, each corresponding to a European Reference Network (ERN), i.e., a network of European centers working on similar rare genetic disorders (Tumiene et al., 2021)) and ii) data analysts organized into working groups of the Data Analysis Task Force (Zurek et al., 2021). Here, we present the systematic analysis performed by the RNA-seq working group of the 521 RNA-seq datasets generated within Solve-RD, comprising 248 undiagnosed families from five disease groups recruited by five ERNs. To manage the complexity of submissions from over 30 contributing centers, we developed a harmonized framework for quality control, outlier detection, and standardized interpretation, supported by collaborative review events (including structured workshops termed “Solvathons” (Yépez et al., 2025)) and shared reporting infrastructure. This framework enabled us to identify pathogenic or likely pathogenic events in 19 out of 248 undiagnosed families, representing a diagnostic rate of 7.7% (Fig. 1).

**Figure 1.**
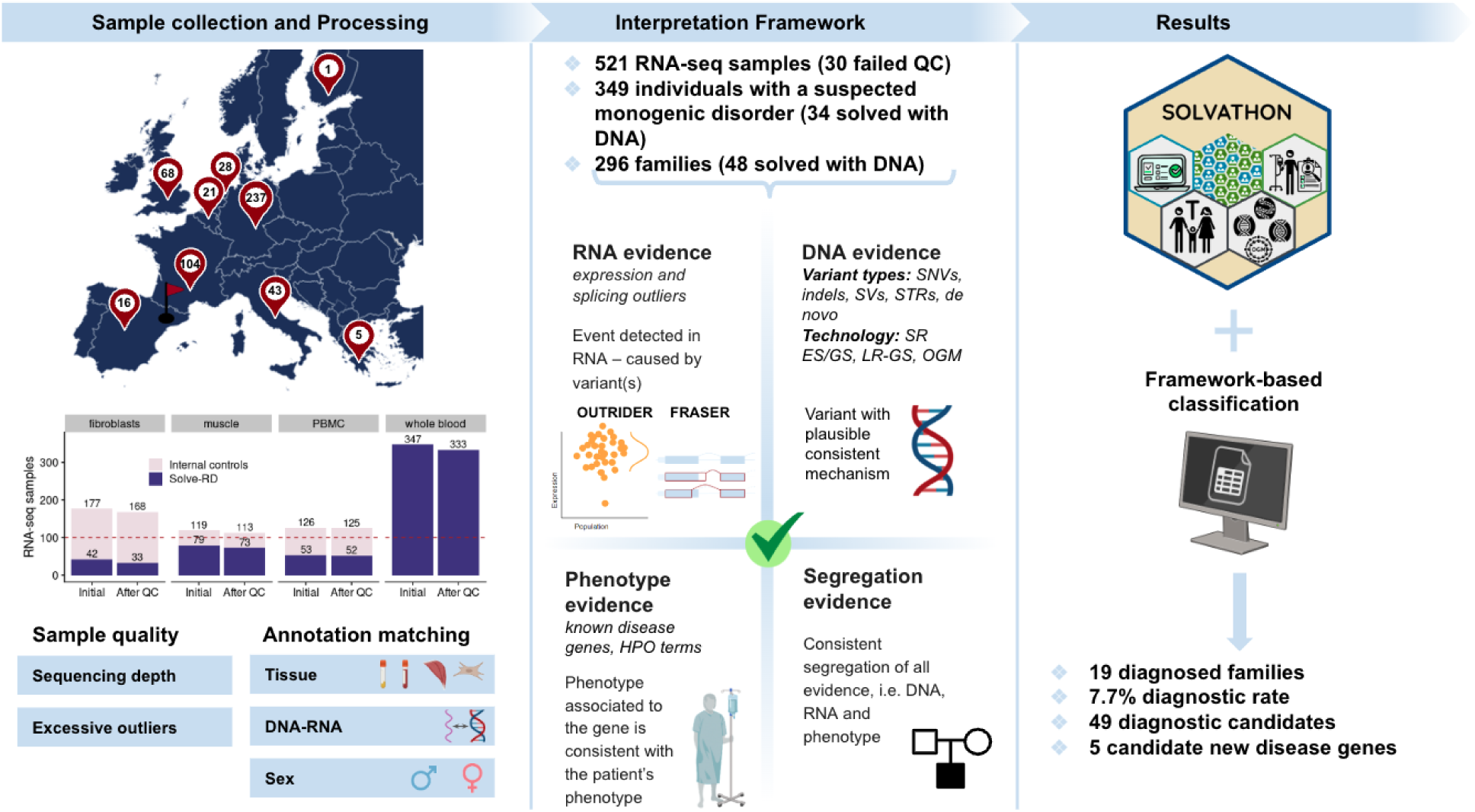
Overview of the Solve-RD RNA-Seq workflow. RNA was extracted from individuals affected by a rare disease and family members (from whole blood, muscle, fibroblasts, and peripheral blood mononuclear cells (PBMC)) and sent to the CNAG in Barcelona, Spain for sequencing. Various quality control steps resulted in some samples being discarded. The RNA-seq data were centrally processed, and the results were integrated with DNA, clinical, and family information. Results were shared with the clinical interpreters via Solvathons (Yépez et al., 2025) and interpreted following a four-step framework. A total of 19 families received a diagnosis.

## Results

### Solve-RD cohort definition and quality control

As part of the Solve-RD consortium, 521 biosamples were collected from undiagnosed rare disease individuals at the start of the project, and in some cases their relatives, and underwent RNA-seq. The affected individuals suffered from neurological, neuromuscular, epilepsy, or immunodeficiency disorders and were selected by expert clinicians from 5 European Reference Networks (ERNs: EpiCARE, EURO-NMD, ITHACA, RITA, and RND), spanning 33 research groups in 9 European countries (Fig. 1, Fig. S1, Fig. S2, Methods). The samples originated from four types of biospecimens: whole blood, skeletal muscle, peripheral blood mononuclear cells (PBMC), and skin-derived fibroblast cultures.

All samples were collected, sequenced, and processed using a unified workflow. The bioinformatics processing was based on STAR for alignment (Dobin et al., 2013) and DROP (Yépez et al., 2021) for feature counting and outlier detection (Methods). Quality control included computational assessment of aligned reads, number of outliers, and consistency of sample metadata regarding sex, tissue, and corresponding DNA (Fig. 2A).

**Figure 2.**
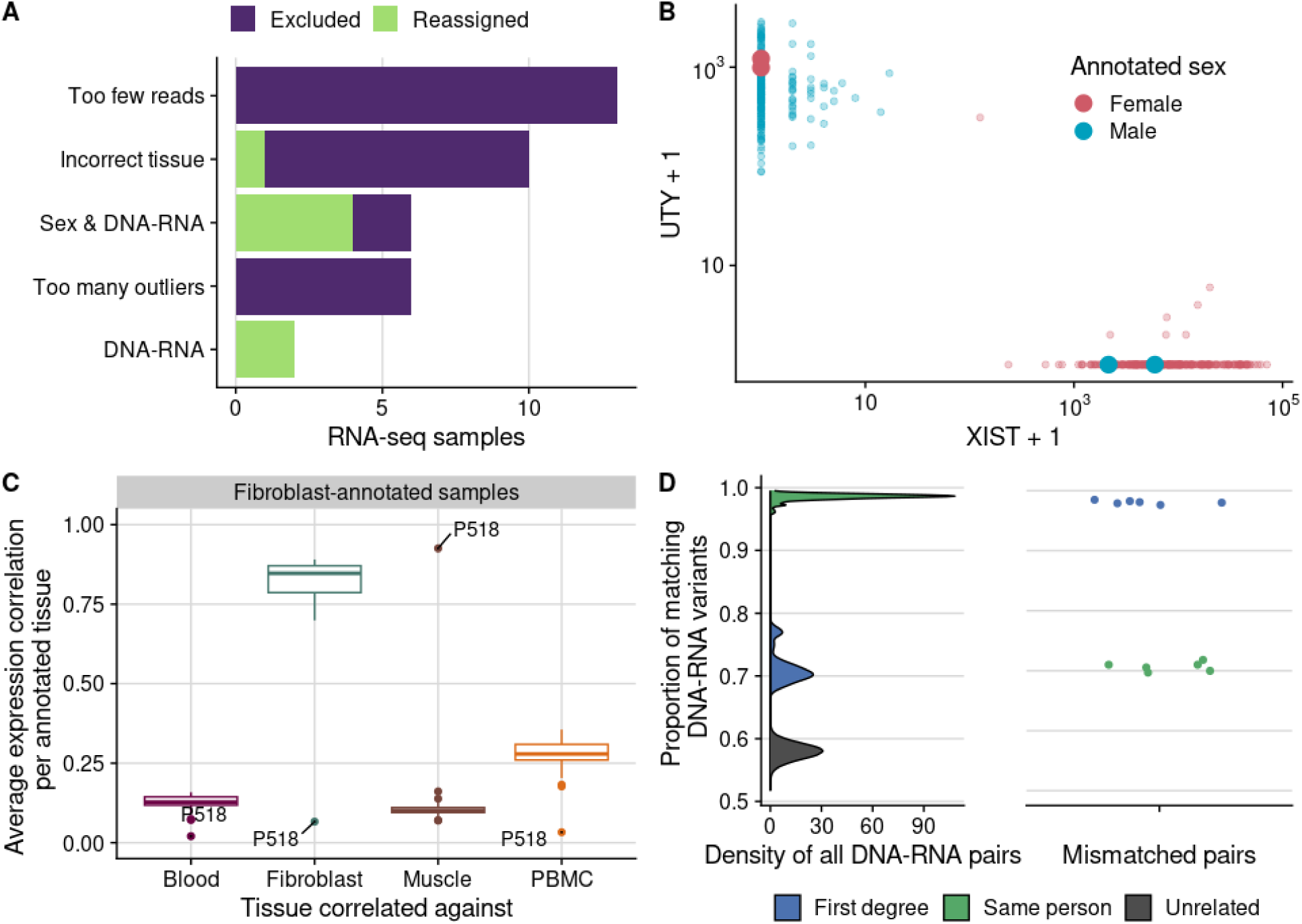
Quality control and sample annotation consistency. **A)** Results of all quality control steps indicating the number of excluded and reassigned samples. Only the reassigned samples are shown in B-D, as those excluded were removed from the cohort altogether. **B)** Gene expression counts aligned to the *XIST* and *UTY* genes, colored by the sex specified in the annotation. Larger dots highlight the four samples with discordant sex annotations, which could be corrected. **C)** Average gene expression correlation (in FPKM) of all 168 fibroblast samples against all the Solve-RD samples, stratified by tissue. One sample, P518, initially annotated as a fibroblast, correlated more strongly with muscle and was subsequently identified as an annotation error. **D)** Density distribution of DNA–RNA single-nucleotide polymorphism matching rates, stratified by same-individual pairs, first-degree relatives, and unrelated samples. On the right, six swaps from three families are shown.

Samples with fewer than 10 million read pairs (after globin-read removal on the blood cohort) were excluded (n=13, Fig. S3). Sex checks based on *XIST* (female-specific) and *UTY* (male-specific) expression identified six inconsistencies (Fig. 2B). Four were confirmed as annotation errors and corrected. The remaining two were excluded because they could not be matched to the corresponding DNA sample and the origin of the inconsistency could not be resolved.

Tissue annotation was investigated by computing the correlation of the fragments per kilobase of transcript per million (FPKM) for each sample against all others within each tissue type. Ten samples showed low correlation with their respective group. One sample (P518) annotated as fibroblast, but matching the muscle correlation profile, was traced to an annotation error, and was corrected. The remaining nine did not correlate with any other tissue type, suggesting a technical issue, and were removed (Fig. 2C, Fig. S4).

DNA–RNA concordance was assessed based on the proportion of matching single-nucleotide polymorphisms. Samples from the same individual showed over 95% concordance, first-degree relatives 65 to 78%, and unrelated pairs 50 to 63%. Eight DNA–RNA pairs were identified as originating from different individuals. Six represented within-family annotation swaps and were corrected, while the remaining two, whose sex annotations were also incorrect, were excluded (Fig. 2D).

Finally, six samples exhibited more than 300 splicing outliers. Although no clear biological feature was shared, these samples were processed in the same biosampling batch as samples discarded due to low RNA Integrity Number or low read counts, suggesting a technical origin. These samples were therefore excluded.

Altogether, seven samples (1.7%) were successfully reassigned to the correct sex, tissue, or DNA sample, highlighting the importance of systematic quality and annotation checks, particularly in large, multi-partner consortia. Another 28 RNA-seq samples failed quality control and were removed. Two additional samples presented annotation errors that could not be resolved and were also excluded, resulting in 491 samples retained for analysis. These samples correspond to 448 individuals, out of which 349 with a suspected monogenic disorder from 296 families, as RNA-seq was performed on multiple tissues for 43 individuals. At the time of inclusion, all individuals were considered genetically undiagnosed. However, during the course of the Solve-RD project, 64 individuals received a diagnosis through DNA re-analysis, leaving 315 samples from 285 individuals across 248 families still awaiting diagnosis.

### Annotation framework allows classification of RNA outliers

The 491 samples that passed quality control were grouped by tissue for outlier detection. Fewer samples than the 100 recommended for outlier detection were available within Solve-RD for fibroblasts (n=33), muscle (n=73), and PBMCs (n=52), and thus were supplemented with RNA-seq samples generated outside the consortium using comparable experimental protocols (Methods). Expression outliers were detected for each sufficiently expressed gene (10,758 in blood, 12,138 in muscle, 12,362 in fibroblasts, and 13,637 in PBMC, Fig. S5) using OUTRIDER (Brechtmann et al., 2018), a denoising autoencoder–based method that corrects for technical and biological variation in RNA-seq read counts and models each gene using a negative binomial distribution. Additionally, aberrant splicing was called at the intron level using FRASER 2.0 (Scheller et al., 2023). We collectively refer to expression outliers and splicing outliers as RNA outliers. Each analysis was performed independently for each tissue type. Across all four tissue sources, coexpression patterns due to both technical and biological effects present in the raw data were no longer visible after normalization (Fig. S6-9).

Restricting the analysis to the 315 samples from previously undiagnosed individuals, we detected a median of three expression outliers (one overexpression, two underexpression), and 5 aberrantly spliced genes per sample (Fig. 3A, Methods). These RNA-seq findings were integrated at the gene level with rare variants identified by short-read exome and genome sequencing, long-read genome sequencing, as well as chromosomal aberrations detected by optical genome mapping, when available. In addition, sample–gene phenotype similarity scores were computed based on HPO terms (Methods). The resulting annotated tables, together with relevant plots, were compiled into per-family reports for downstream interpretation. For the 20 samples presenting more than 20 splicing outliers and two with more than 20 underexpression outliers, we retained only the top 20 of each based on genetic and phenotypic relevance (Fig. 3A, Methods). Overall, this filtering resulted in 3,004 outliers to be interpreted across the cohort (Fig. 3B). Genes that were both expression and splicing outliers in one sample were interpreted only once.

**Figure 3.**
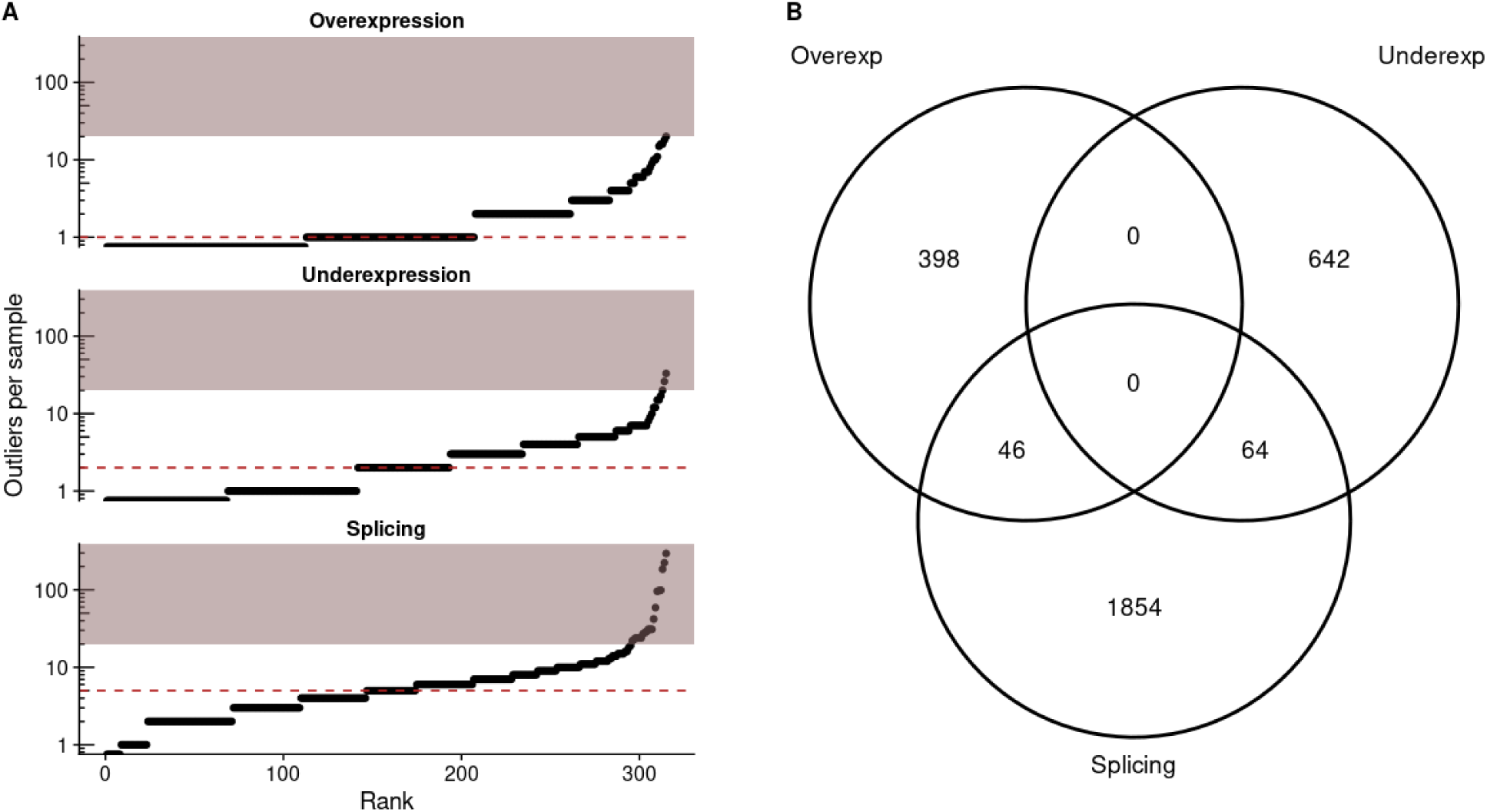
Expression and splicing outliers. **A)** Distribution of overexpression, underexpression, and splicing outliers in the 315 samples from previously undiagnosed individuals. The horizontal dashed line denotes the median. For samples with more than 20 outliers (highlighted area), only those either genetically or clinically relevant were retained. **B)** Overlap between the different categories of outliers. Each value represents a sample-gene combination. To ensure coherent interpretation of RNA-seq findings across the Solve-RD consortium, we established a standardized annotation framework, combining evidence from four aspects: 1) RNA outlier reliability, 2) phenotypic similarity of the patient to the phenotype previously described for patients with disruptions in this gene, 3) consistency of the DNA variant mechanism with the detected RNA outlier, and 4) RNA outlier, DNA variant, and clinical segregation within the family (Fig. 4A). Each of the four classes of evidence was divided into categories by strength, which when combined, yielded an overall evidence level classification ranging from “discarded” to “very strong evidence” (Methods, Fig. 4A). For the variant, phenotype, and segregation assessment, we used the ACMG guidelines and the ClinGen SVI Splicing Subgroup recommendations (Richards et al., 2015; Walker et al., 2023) and added how to interpret them in the context of the RNA outliers (Methods).

In total, 3,004 RNA outliers were evaluated for potential pathogenicity using this framework. The vast majority (2,519; 84%) were discarded, most commonly due to insufficient phenotypic concordance (2,146), followed by unreliable RNA outliers (146) and lack of consistently segregating variant support (132; Fig. 4B). A further 368 RNA outliers were classified as functionally unclear. These were supported by a genetic variant in a gene already associated with disease and were potentially consistent with the patient’s clinical presentation, but lacked sufficient functional evidence.

**Figure 4.**
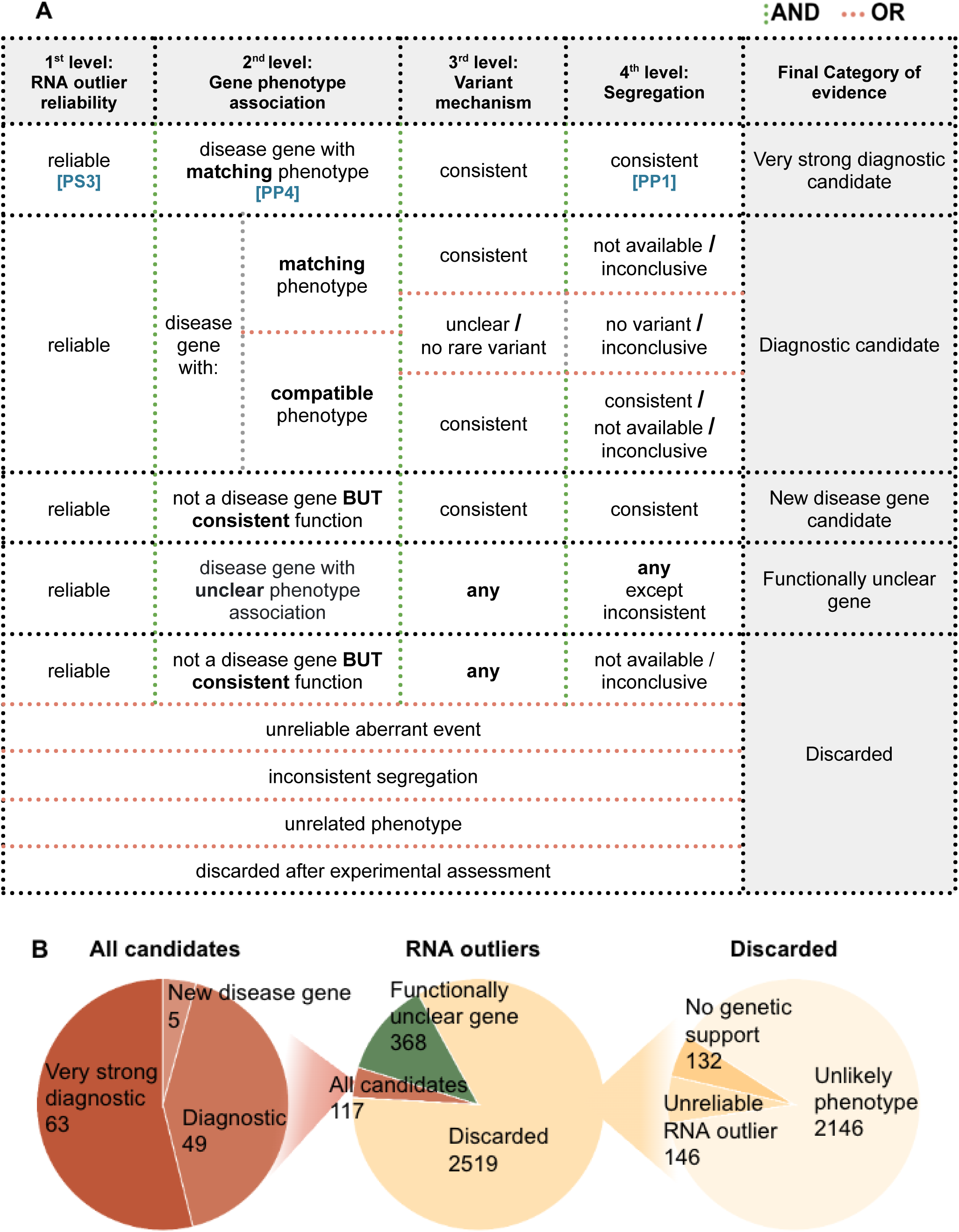
Results classification framework. **A)** RNA outlier classification framework built on four evidence levels with different resulting pathogenicity outcomes. In the case of very strong diagnostic candidate, RNA-seq support can be considered as a positive functional study, i.e., pathogenic strong evidence criterion 3 (PS3), genotype–phenotype association to pathogenic supporting evidence 4 (PP4), and consistent segregation to pathogenic supporting evidence 1 (PP1) as defined by the ACMG guidelines (Richards et al., 2015). **B)** Pie charts showing the outcomes of the RNA outlier interpretation framework, from left to right: candidate outliers, overall RNA outlier classification, and exclusion criteria of discarded cases.

Using this framework, 117 RNA outliers (4%) were prioritized as candidates for diagnosis (Fig. 4C). Among these, 63 were classified as very strong diagnostic candidates, 49 as diagnostic candidates lacking one line of strongest supporting evidence (genetic, clinical, or segregation), and 5 as potential novel disease genes. The 63 very strong candidates were confirmed as pathogenic by the referring clinicians and corresponded to 21 diagnosed individuals from 19 families, including three patients from the same family. In three patients (P07, P20, P21), large deletions or duplications resulted in 21, 9, and 15 expression outlier genes, respectively. The remaining 18 patients had monogenic diagnoses.

This annotation framework ensured comparable decisions across interpreters. It also allowed us to record annotation outcomes at intermediate investigation stages and facilitated revisiting specific categories when new clinical, genomic (such as matched long-read genome sequencing), or segregation data became available, to conclude on the final interpretation of outliers in the RNA-first approach.

### RNA-seq analyses resolved a broad spectrum of pathogenic variant mechanisms

Aberrant expression and splicing analyses directly contributed to the diagnosis of 21 individuals from 19 families, representing a diagnostic rate of 7.7% of the 248 undiagnosed families (Fig. 5, Table 1). Consistent RNA-seq evidence was found for a broad range of pathogenic genetic variant types, including SNVs and short indels in exonic, splice-region, and deep intronic positions (n = 12); structural variants such as deletions, duplications, and translocations (n = 6); and short repeat expansions (n = 2). The total number of variants was 20, because one patient (P19) showed compound heterozygosity involving a structural variant and a short indel. In ten of the newly solved families, the initial evidence for diagnosis was a DNA-based assay, i.e., candidate variant but missing functional evidence. In contrast, in nine families RNA-seq unravelled the disease-causing gene, highlighting the added value of the RNA-first approach (Fig. 5, Table 1).

**Figure 5.**
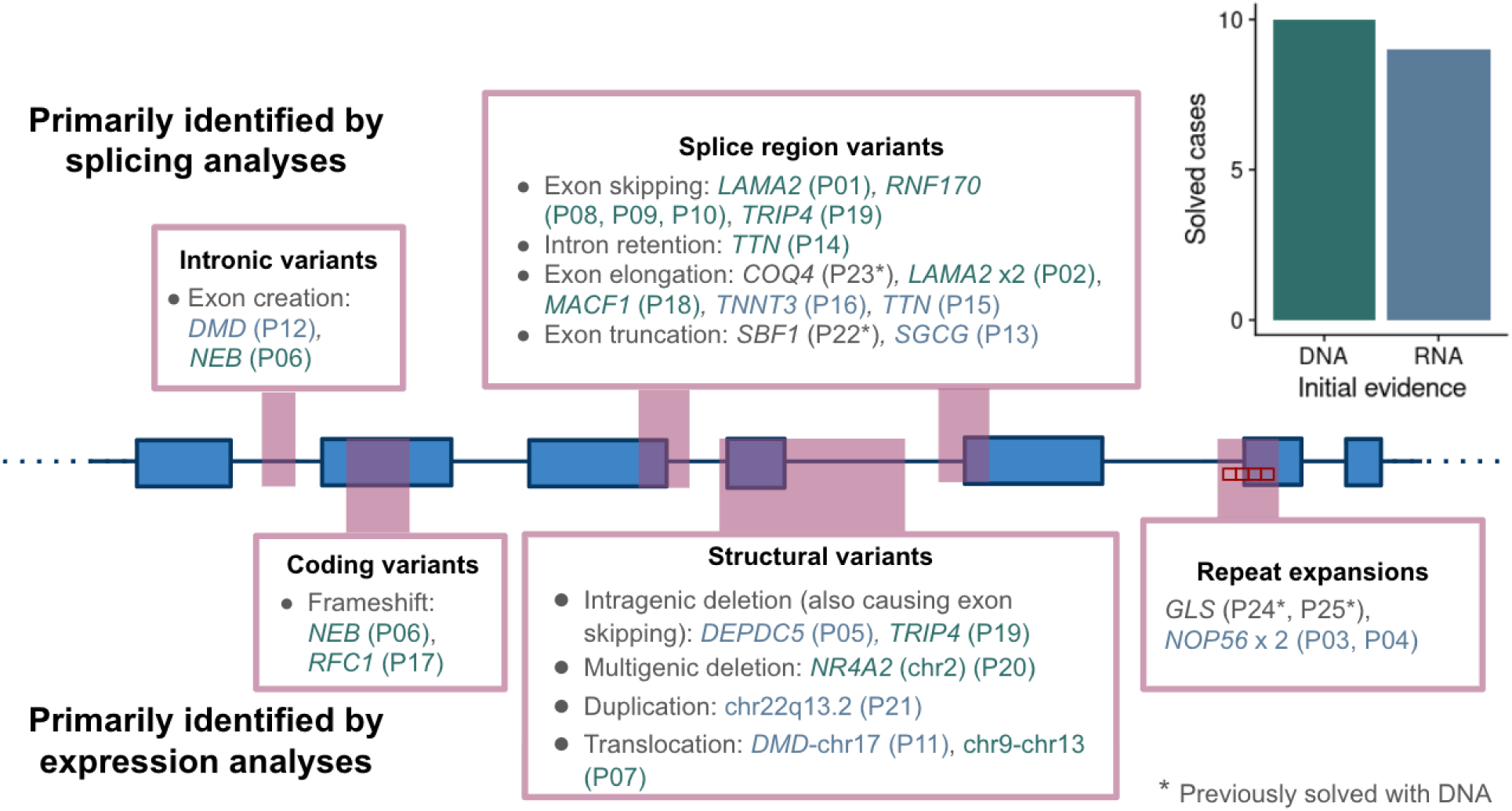
Overview of solved cases. Schematic gene with introns (lines) and exons (boxes) showing the solved cases grouped by type of causal variant and whether they were primarily detected through expression or splicing analyses. Genes with a star represent diagnoses independently reached by genome analysis alone and for which RNA-seq further corroborated the mechanism. *DEPDC5* and *TRIP4* were confirmed by both expression and splicing analyses. On the top right, the number of families for which the initial evidence for diagnostics was DNA (n=10) or RNA (n=9) testing (excluding P22, P23, P24 as they were diagnosed using DNA only). In compound heterozygous cases where one variant had already been identified by DNA analyses and the second variant was unknown, the initial evidence was denoted as DNA.

**Table 1.**
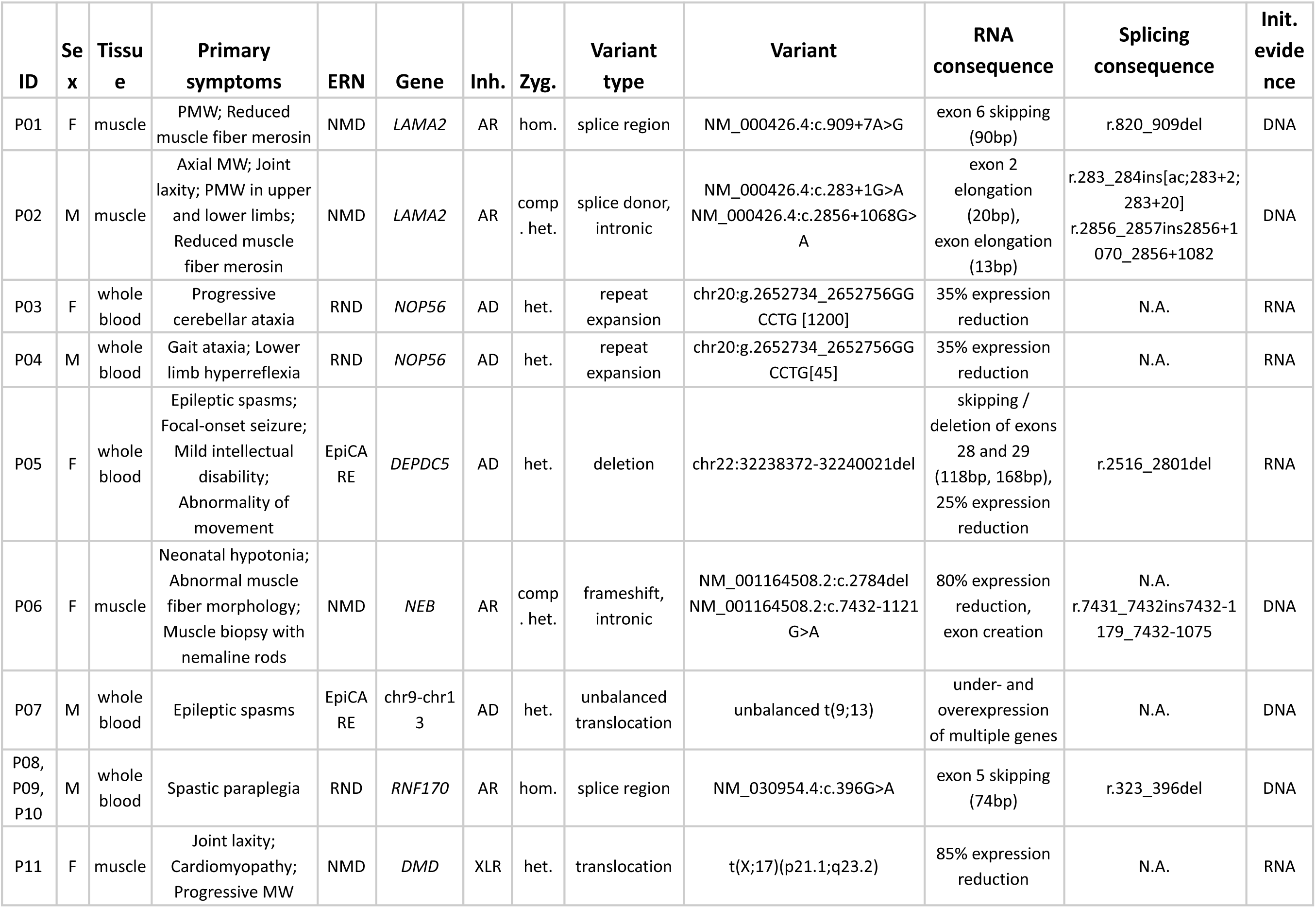
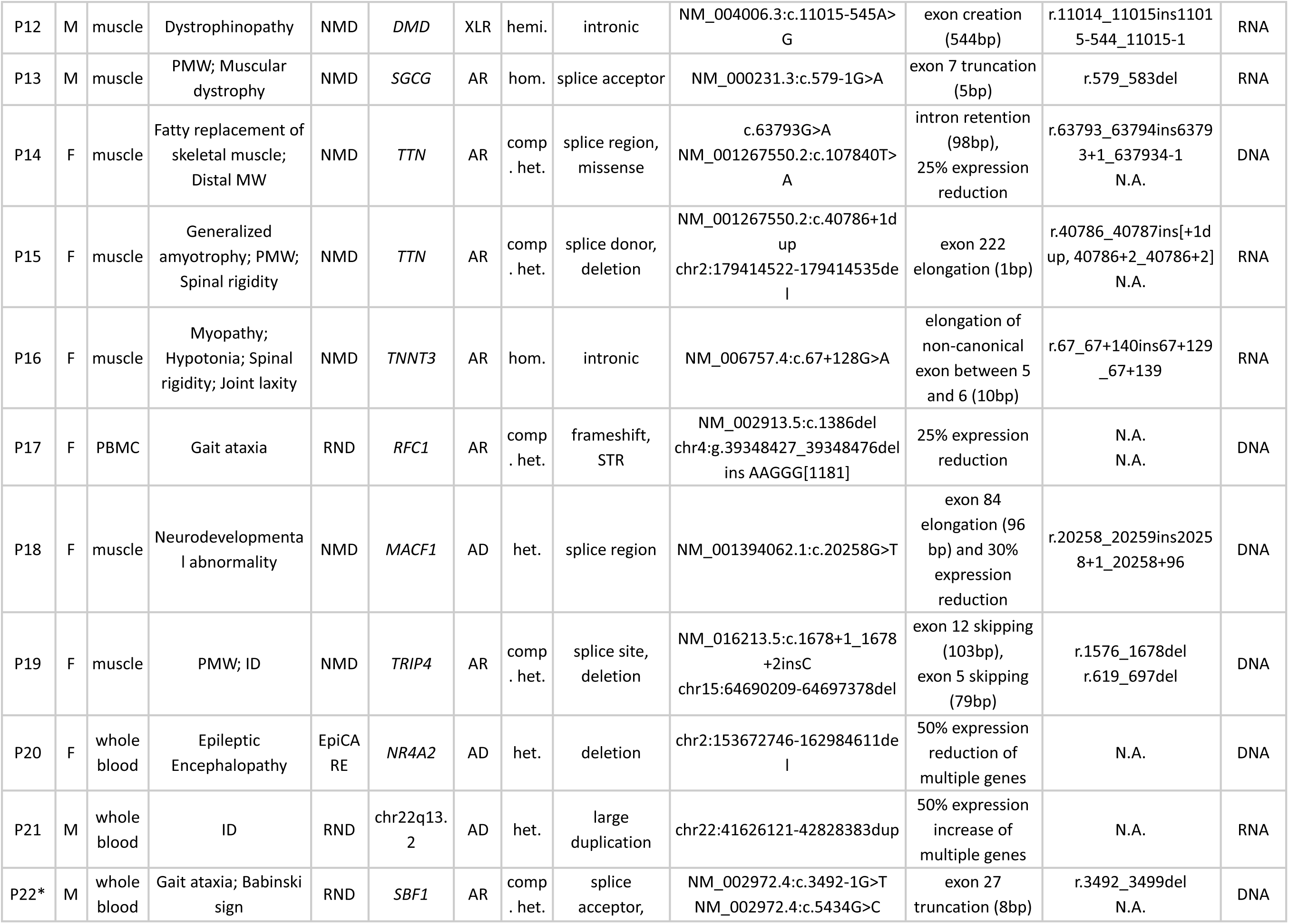
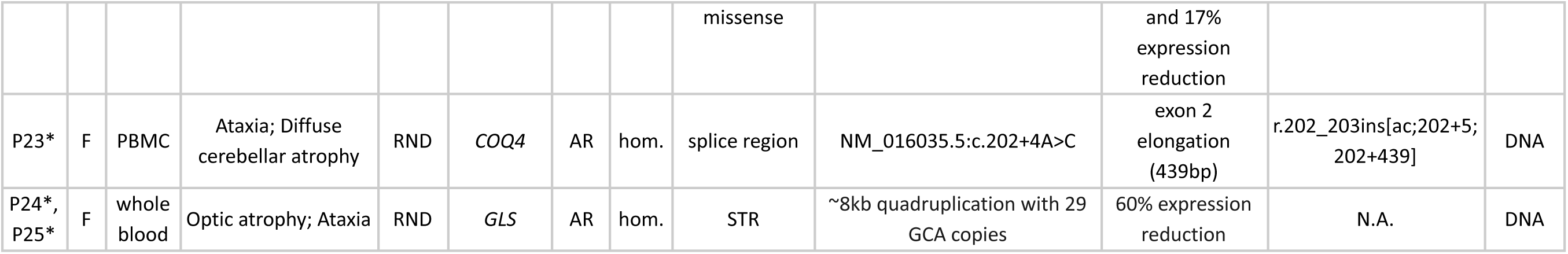
Genetic and clinical overview of the cases diagnosed via RNA-seq. Each row denotes a diagnosed family. The splicing consequence denotes the deletion or insertion as a result of a DNA variant, as suggested by the HGVS Nomenclature (Hart et al., 2024; HGVS Variant Nomenclature Committee, 2025). Variants and splicing consequences are reported with respect to the MANE Select transcript (Morales et al., 2022). Patients P22-P25 had already been diagnosed with DNA analyses and are highlighted with a star. Zyg.: zygosity, Inh.: inheritance, Init.: initial, F: female, M: male, AD: autosomal dominant, AR: autosomal recessive, XLR: X-linked recessive. (P)MW: (proximal) muscle weakness, ID: intellectual disability. N.A.: not applicable. Six patients (P01 - LAMA2, P06 - NEB, P11 - DMD, P12 - DMD, P13 - SGCG, and P16 - TNNT3) have already been reported in disease-specific studies (Estévez-Arias et al., 2024; Neuhoff et al., 2024; Segarra-Casas et al., 2024; Müller et al., 2025), and three in the main Solve-RD LR-GS study (P03 and P04 - NOP56, P17 - RFC1, Steyaert et al, 2024)).

In addition, three families (P22 - P25) were independently solved using genome analyses only (Cordts et al., 2022; Fazal et al., 2023) and subsequently confirmed using RNA-seq. Two of these families harbored SNVs resulting in exon elongation and exon truncation, and the other one a repeat expansion in a gene with aberrant expression. Together, these results underscore the wide spectrum of pathogenic variations that can be revealed or corroborated through expression and splicing analyses, highlighting their value as complementary tools in rare disease diagnostics.

### Expression and splicing outliers in genetically solved individuals

The fold changes computed by OUTRIDER reflect the relative gene expression levels for each individual compared to the cohort average after having adjusted for sources of variation shared across genes, which may be biological (e.g. sex, age) or technical (e.g. sequencer, batch), in an unsupervised manner (Brechtmann et al., 2018; Yépez et al., 2022). In this cohort, variants predicted to trigger nonsense-mediated decay (NMD) on both alleles were accompanied by expression reductions greater than 70% in two solved cases: a female patient (P11) with a heterozygous translocation in *DMD* and a female patient (P06) with compound heterozygous variants in *NEB* (frameshift plus an intronic variant generating a novel exon with a premature stop codon). Variants predicted to trigger NMD on a single allele were accompanied by expression reductions of approximately 25 to 40% in three solved cases: intragenic deletions in *TRIP4* and *DEPDC5*, and a frameshift variant in *RFC1* (Fig. 6A).

**Figure 6.**
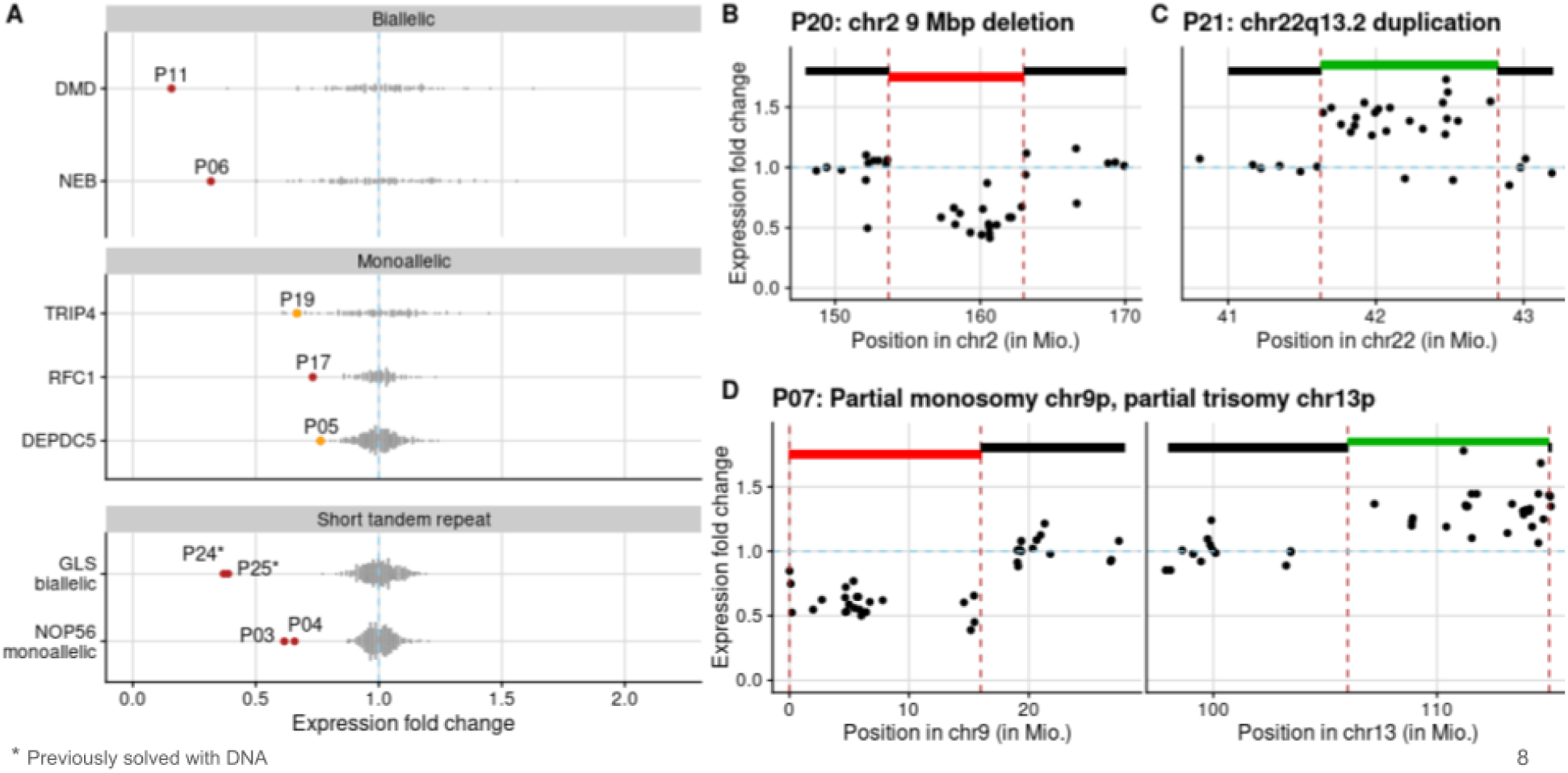
Expression outliers. **A)** Distribution of the expression fold change of the 7 genes for which aberrant expression was found in the disease-causing gene stratified by biallelic and monoallelic NMD-triggering variants, and short tandem repeats. Each dot represents one sample. **B-D)** Distribution of the expression fold change of individual P20 harboring a 9 Mbp deletion (B), P21 harboring a duplication in 22q13.2 (C), and P07 harboring an unbalanced translocation with partial monosomy and partial trisomy (D). Each dot represents a gene. Deletions are marked in red; duplications in green.

Aberrant expression was also observed in four individuals from three families with pathogenic repeat expansions. In two individuals from the same consanguineous family, the repeat expansion was present on both alleles of exon 1 of *GLS* (Fazal et al., 2023), accompanied by a decrease in expression of 60% and 62%. In the remaining two unrelated individuals, the expansion affected only one allele in intron 1 of *NOP56* (at the known locus associated with spinocerebellar ataxia type 36, SCA36 (Kobayashi et al., 2011)), exhibiting 33% and 35% reduced expression. These findings are consistent with transcriptional silencing mechanisms reported for other repeat-expansion-associated disorders (Groh et al., 2014; Malik et al., 2021). In addition, P17 also harbored a pathogenic repeat expansion on one allele of *RFC1* together with a heterozygous frameshift variant on the other allele. A 25% expression reduction was observed in that gene; however, it is presumably due to the frameshift variant, as its allelic ratio was 26% (36 reads containing the frameshift from a total of 139 reads at that position).

Multigene deletions and duplications are reflected in expression data as contiguous blocks of underexpressed or overexpressed genes, as observed in three individuals from this cohort (Fig. S10). In the first, a 9 Mb deletion on chromosome 2 was associated with a 40 to 60% reduction in expression across nearly all genes within the affected region (Fig. 6B). In the second patient, a 1.1 Mb duplication at 22q13 exhibited a 30 to 60% increase in gene expression in 18 genes located in that region (Fig. 6C). Finally, in the third, an unbalanced translocation with partial monosomy of chromosome 9 and partial trisomy of chromosome 13 was associated with approximately 50% decreased expression in the monosomic segment and 50% increased expression in the trisomic segment (Fig. 6D). In all three cases, the aberrant expression pattern confirmed the genetic alteration found by DNA analyses.

These examples illustrate the power of aberrant expression analysis for identifying copy number variants and other complex structural rearrangements, as well as short variants that lead to NMD-mediated transcript depletion. They also highlight its potential utility for detecting disease-related repeat expansions.

Aberrant splicing analyses were also instrumental in achieving new diagnoses. Variants affecting splicing can result in either insertions (e.g. exon elongation, exon creation, or full intron retention) or deletions (e.g. exon skipping or exon truncation) in the RNA sequence. In this cohort, we identified variants located in the splice region (i.e., within 1-3 bp inside an exon or 1-8 bp inside the intron adjacent to a splice site) that potentially disrupted the canonical donor or acceptor sites. These led to exon elongation (n=5), exon truncation (n=2), exon skipping (n=3), and one full intron retention. In addition, we observed a variant in the 10^th^ bp upstream of the acceptor site of an exon located between exons 5 and 6 of the MANE Select transcript of *TNNT3* (P16) that created a novel acceptor site, reflected as a 12 bp exon elongation; two rare deep intronic variants likely activating the creation of new exons; and two intragenic deletions manifesting as exon skipping events (Fig. 7A).

**Figure 7.**
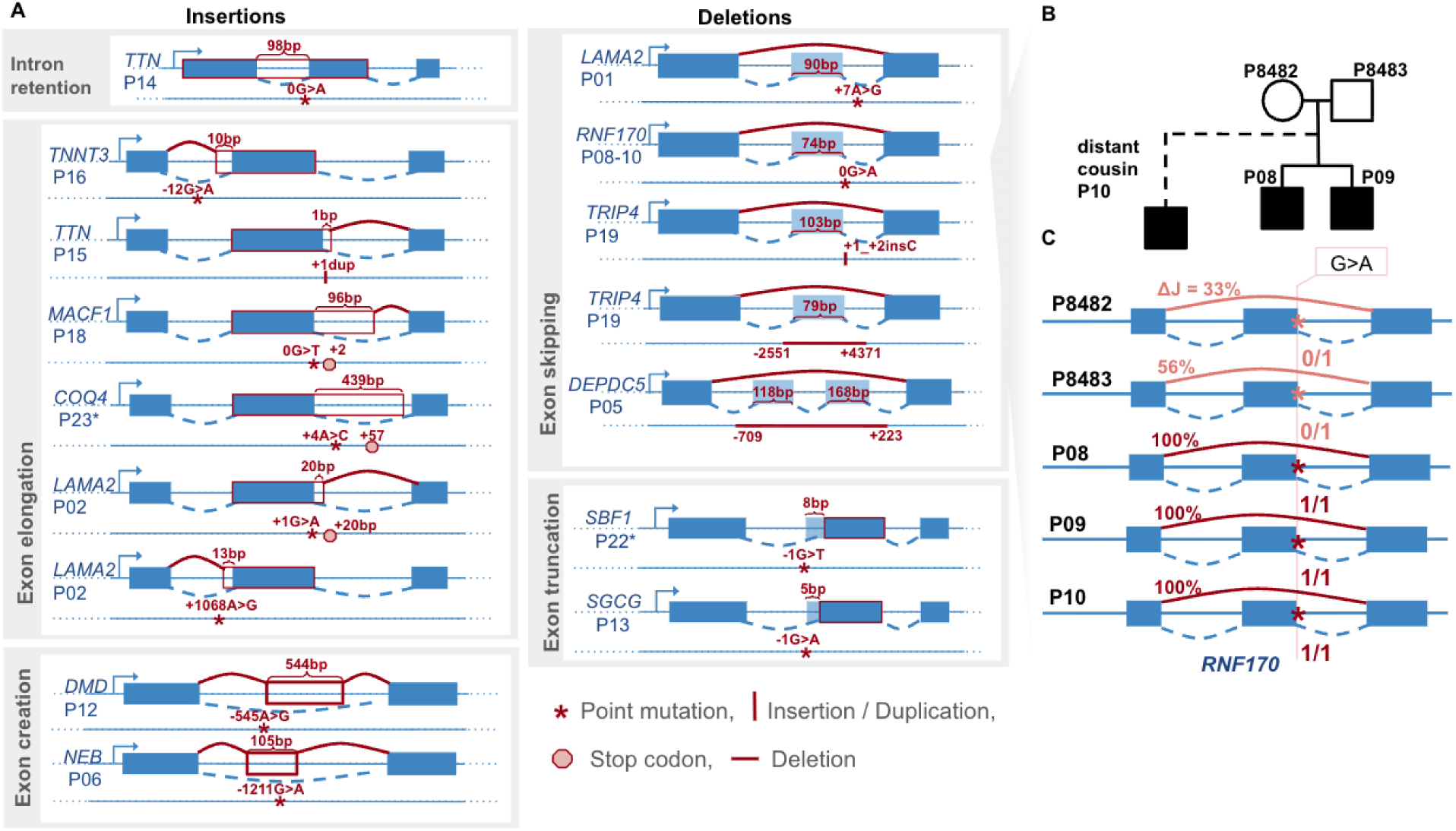
Splicing outliers. **A)** Depiction of splicing outliers split into groups depending on their type. Type and location of causal variant, location of the stop codon, and length of insertion or deletion are also indicated. **B)** Pedigree of a family carrying a splice-region variant in the *RNF170* gene. **C)** Schematic of the exon-skipping event observed in the family shown in panel (B), associated with a variant at the last exonic bp of the donor site of exon 5. The percentages correspond to the differential in splicing, reflecting that heterozygous carriers (parents) exhibit ∼50% exon skipping, whereas homozygous individuals show nearly complete exon skipping.

### RNA-seq co-segregation

For 69 individuals, we also collected RNA-seq samples from their relatives, allowing us to assess the segregation of the aberrant RNA-seq events with the phenotype and with the suspected causal variants. In one such family, RNA-seq was performed for both unaffected parents, two affected children, and one affected distant cousin (Fig. 7B). Splicing analysis revealed skipping of the fifth exon of *RNF170*. The parents, who carried a heterozygous SNV at the last base of the skipped exon, showed partial exon skipping (differential splicing, delta Intronic Jaccard Index, ΔJ, of 33% and 56%). In contrast, the two affected children and the affected cousin, who were homozygous for the same variant, showed complete exon skipping, supporting consistent segregation of the RNA defect within the family (Fig. 7C).

## Discussion

Here we report a standardized workflow and results derived by deploying RNA-seq-based rare disease clinical research across a pan-continental consortium that is heterogeneous with respect to disease entities, collected biomaterials, and the degree of expertise with RNA-seq data across the involved partners. We establish a standardized interpretation framework for RNA-seq results that is compatible with the ACMG diagnostic guidelines. The framework supports consistent classification of the findings across the 33 sample-submitting groups. Close collaboration between data analysts and clinical interpreters facilitated by two in-person Solvathon workshops (Yépez et al., 2025) and detailed integrative reports substantially contributed to resolving these very complex cases and increased the RNA-seq analysis expertise across the consortium. In addition, Solve-RD’s working group structure facilitated efficient access to preprocessed genetic and clinical data, which further streamlined the interpretation process. This effort resulted in molecular diagnoses for 19 out of 248 undiagnosed families, i.e., 7.7% additional diagnostic yield, and confirmed three families previously resolved with DNA analysis.

Data interpretation is inherently dynamic due to updates in family genetic and clinical data, as well as scientific advances reflected in updated clinical variant relevance and disease gene entries, e.g. OMIM (Amberger et al., 2019) or ClinGen (Andersen et al., 2025). The classification framework presented here provides a coherent and unified way of not only annotating, but also updating RNA-seq interpretation. For example, P11 with a *DMD* translocation was initially classified as a *Diagnostic candidate* due to reliable RNA outlier in a disease gene with matching phenotype, but no rare variant was found in the exome sequencing data. The translocation was identified months later, only after performing and analyzing short-read genome sequencing data (Segarra-Casas et al., 2024). Similarly, several of the other different types of candidates found here could be upgraded after reanalysis of available data or generation of new data, for instance, by long-read sequencing or optical genome mapping. Hence, having a consistent framework for recording intermediate interpretations facilitates revisiting the cases, guides additional analyses with a focused search for genetic variants in candidate genes, and enhances collaborative work on the cases.

A molecular diagnosis provides opportunities for targeted treatment and informed genetic counseling (Pogue et al., 2018). Targeted antisense oligonucleotide (ASO) treatment can correct aberrant splicing at specific loci (Kim et al., 2019; Kumar et al., 2022). Following the guidelines recently reported by (Zardetto et al., 2024; Cheerie et al., 2025), only variants beyond ±15 bp of a junction could be potential candidates for this treatment. In our cohort, three patients (P06, P12, and P16) had such deep intronic variants. However, all of them affect the muscle, which is a tissue that currently cannot be sufficiently well targeted by this technology (Roberts et al., 2020). In this cohort, reaching a diagnosis enabled appropriate antiseizure therapy for patient P20 and offered several families guidance for counseling and reproductive planning (Table S1).

The tissue of origin of the biosamples has a drastic effect on the genes that can be analysed. Fibroblasts, PBMCs, whole blood, and muscle biopsies each express overlapping, yet distinct and jointly comprehensive, sets of disease-relevant genes. Across the five principal disease groups studied (corresponding to the five ERNs), blood exhibited the lowest number of expressed genes, with the exception of immunological disorders, consistent with previous reports (Aicher et al., 2020; Murdock et al., 2021; Yépez et al., 2022). In contrast, fibroblasts captured the largest number of expressed genes in most disease groups, except for neuromuscular disorders, where muscle showed the highest gene coverage, and immunological disorders, where PBMCs captured the most expressed genes (Fig. S2). Transdifferentiating fibroblasts into neurons has shown to result in a higher number of genes expressed than in the original fibroblasts (Li et al., 2024; Nicolas-Martinez et al., 2024). However, secondary alterations during the reprogramming and differentiation processes can introduce additional noise.

RNA-seq can reveal complex patterns of transcriptional dysregulation. Samples with unusually high numbers of splicing or expression outliers may result from technical or ancestry-related effects (Yépez et al., 2022), but can also reflect biological mechanisms. These include defects in spliceosome components (such as *SON* in ZTTK Syndrome (Kim et al., 2016) and those seemingly caused by pathogenic variants in the small nuclear RNAs *RNU4-2* (Chen et al., 2024; Nava et al., 2025), *RNU4ATAC* (Arriaga et al., 2025), and *RNU2-2* (Jackson et al., 2025)); downstream regulatory effects (e.g. mitochondrial gene downregulation due to *LIG3* or *LRPPRC* (Yépez et al., 2022), or histone gene upregulation due to *RNU7-1* (Chui et al., 2025)); or large deletions and duplications covering multiple genes (such as those presented in P07, P20, and P21 Fig. 6B-D). Consequently, RNA outliers should also be interpreted in the context of variants in other genes, to avoid false pathogenicity assignments and to distinguish true biological signals from artefacts. In this cohort, none of the samples with more than 80 splicing outliers had pathogenic variants in neither of those snRNAs.

Overall, this multi-center study spanning multiple diseases and tissues reinforces the value of RNA-seq as a complement to DNA sequencing for rare disease diagnosis. The framework presented here provides a starting point for future diagnostic guidelines and can be further extended, generalized, and optimized. It will serve as the backbone for RNA-seq analyses within the ERDERA project (www.erdera.org) and could be applied to other large-scale initiatives, such as GREGoR (Dawood et al., 2025). More diverse tissues including in-vitro differentiated neurons, deeper sequencing, transcriptome-wide regulatory signatures, long-read RNA technologies, and multi-omics data layers can further improve the diagnostic rate and the understanding of diseases. In addition, reproducibility and robustness studies are needed to aim for establishing RNA-seq in routine practice together with DNA sequencing, such as that recently undertaken by (Zhao et al., 2025). Moreover, with larger datasets with matched genomes and transcriptomes across a broad set of diseases, and a large number of diagnostics, the setting of significance cutoffs could be revisited, for instance using the odds of pathogenicity approach (Tavtigian et al., 2018; Smirnov et al., 2022). Ultimately, continued advances in therapeutic development and clinical care will be essential for translating these molecular insights into meaningful outcomes, including therapies, for affected individuals.

## Material and Methods

### Solve-RD RNA-seq cohort

Individuals suspected to suffer from a rare genetic disorder and with inconclusive DNA evaluation were recruited within Solve-RD for RNA-seq analysis as part of the combined-omics approach strategy (Zurek et al., 2021). All individuals were recruited via a European Reference Network (ERN). For this study, five ERNs contributed with samples: RND focusing on rare neurological diseases; EURO-NMD studying neuromuscular disorders; ITHACA focusing on rare congenital malformation syndromes and intellectual disability; EpiCARE focusing on rare and complex epilepsies; and RITA investigating rare immunodeficiency, autoinflammatory, and autoimmune diseases. No individual was excluded based on sex, gender, ethnicity, race, age, or any other socially relevant groupings. In addition, each ERN shared a list of genes known to be associated with the disease it studies (Laurie et al., 2025), which we used to prioritize the results.

### RNA extraction, storage and shipment

Total RNA for short-read RNA-seq was extracted at the local clinical or research laboratories, following the consortium standard operating procedures (SOPs) based on manufacturer best practices (Supplementary information). RNA extraction was not centrally monitored. Local sites were allowed to use equivalent protocols, provided they met the Solve-RD input and quality requirements described below.

For whole blood, venous blood was collected into PAXgene Blood RNA tubes (2.5 mL, PreAnalytiX / BD, cat. no. 762165), which were drawn as the last tube in the venipuncture procedure. Immediately after collection, tubes were gently inverted 8–10 times and kept upright at room temperature (18–25°C) for 2-72 h. Tubes were then frozen at −20°C for 1-7 days and subsequently stored at −80°C until RNA isolation. RNA was extracted using the PAXgene Blood RNA Kit (Qiagen / PreAnalytiX, cat. no. 762174) according to the manufacturer’s instructions and the Solve-RD “RNA isolation from PAXgene tubes” SOP.

For fibroblasts and muscle, RNA isolation followed the Solve-RD SOP. Samples were processed on ice as rapidly as possible after collection. Tissue or cell pellets were either snap-frozen in liquid nitrogen and stored at −80°C or immediately lysed in guanidinium-based lysis buffer (e.g. RLT) containing RNA stabilizer. The RNAlater cryoprotectant was used according to the manufacturer’s instructions. Total RNA was isolated using Qiagen RNeasy micro or mini spin-column kits, selected according to input amount, following the manufacturer’s protocol. Homogenization was performed using bead-based lyser systems, rotor–stator devices or shredder columns, depending on tissue type and local infrastructure. On-column DNase digestion was applied to minimize genomic DNA contamination.

Isolated RNA was eluted in a nuclease-free buffer, aliquoted into RNase-free 1.5 mL LoBind tubes and stored at −80°C until shipment. Consortium guidelines requested shipment of ≥1–2 µg total RNA with RNA integrity number >7 on dry ice to the central hub at the Radboudumc, Nijmegen, The Netherlands, and from there to the sequencing center (CNAG, Centro Nacional de Análisis Genómico, Barcelona, Spain). At Radboudumc, RNA concentration and integrity were measured prior to redistribution, and only samples that met the quantity and quality requirements were forwarded.

### RNA library preparation and sequencing

At CNAG, samples were received in 7 batches. Total RNA samples underwent an independent quality control using the Qubit RNA BR Assay kit (Thermo Fisher Scientific) to assess quantity and the Agilent Fragment Analyzer DNF-471 RNA Kit (15 nt) to evaluate integrity. RNA-seq libraries were generated from total RNA using the TruSeq Stranded mRNA Library Prep Kit (Illumina). Polyadenylated RNAs were enriched with oligo-dT magnetic beads from 500 ng of total RNA. The resulting blunt-ended double-stranded cDNA was 3’ adenylated, and Illumina platform-compatible adaptors with unique dual indexes and unique molecular identifiers (Integrated DNA Technologies) were ligated. The ligation product underwent enrichment with 15 PCR cycles. The final library was validated using an Agilent Bioanalyzer DNA 7500 assay. Libraries were sequenced on an Illumina NovaSeq 6000 instrument in paired-end mode with a read length of 2x151 bp, following the manufacturer’s protocol for dual indexing. Image analysis, base calling, and quality scoring were processed using the manufacturer’s software Real Time Analysis v.3.4.4, followed by the generation of FASTQ sequence files.

### RNA-seq data processing and quality controls

RNA-seq reads were trimmed with Trim Galore v0.6.7 (Krueger, 2021) and mapped against the hg19 human reference with STAR v.2.7.8a (Dobin et al., 2013) using the two-pass mode for each sample separately (--twopassMode = BASIC). Mapping quality metrics were calculated with Qualimap (García-Alcalde et al., 2012), featureCounts (Liao et al., 2019), and STAR. The aberrantExpression module DROP v1.3.3 was used to verify sex assignment (Yépez et al., 2021). Matching of the annotated tissue was performed by computing the correlation of the log-centered size factor-normalized counts and taking the average of each sample against the rest of the samples of each of the four tissues. The tissue with the highest average correlation was assigned as the computationally predicted tissue.

### RNA-seq data analysis

#### Detection of aberrant expression

Detection of aberrant expression was fully based on DROP v1.3.3 (Yépez et al., 2021). We used as a reference genome the release 34 of the GRCh37 primary assembly from the GENCODE project (Frankish et al., 2021). Read pairs that fell completely within exonic regions were counted using the summarizeOverlaps function from the GenomicAlignments (Lawrence et al., 2013) R package. Reads that fully overlapped more than one feature were assigned to each of those features. For robustness of the statistical fit, genes with a 95^th^ percentile FPKM lower than 1 were considered to be insufficiently expressed for expression outlier analysis and filtered out.

Expression outliers were detected using OUTRIDER (Brechtmann et al., 2018). Significant events were defined as those with an FDR lower than or equal to 0.1 computed across i) all expressed genes, or ii) genes in the corresponding ERN disease gene list, or iii) genes harboring a rare stop, frameshift, or splice region variant. This strategy allowed focusing on the outliers with strongest effect and supported by either genotypic or phenotypic evidence.

#### Detection of aberrant splicing

Detection of aberrant splicing was fully based on DROP v1.4.0, which uses FRASER 2.0 (Scheller et al., 2023), an annotation-free aberrant splicing detection algorithm. The default cut-offs for filtering and significance were used. Exon-exon and exon-intron junctions with less than 20 reads in all samples and for which the total number of reads at the donor and acceptor splice site is 0 in more than 75% of the samples were filtered out. Splicing outlier genes were defined as those in which Holm’s adjusted p-value across junctions of the tested gene was less than or equal to 0.1. Outlier junctions are defined as those in splicing outlier genes, with an FDR less than or equal to 0.1 (also computed on all genes and on the disease genes provided by each ERN) and an effect size (absolute delta intronic Jaccard index) larger than 0.15.

To increase statistical power in both aberrant expression and splicing detection, non Solve-RD RNA-seq samples extracted from the corresponding tissue were added for the fibroblast (n = 135, downloaded from Zenodo (Yepez et al., 2022)), muscle (n = 40, from the August Pi i Sunyer Biomedical Research Institute, IDIBAPS), and PBMC cohorts (n = 73, from the Bellvitge Biomedical Research Institute, IDIBELL). The muscle and PBMC samples were also sequenced at the CNAG following the same protocol described above. The fibroblast samples were sequenced and preprocessed as described in (Yépez et al., 2022). Samples with more than 300 splicing outliers were removed.

### Variant annotation

Structural variants (SVs) and copy number variations (CNVs) in short-read exome sequencing data and short-read genome sequencing data were called using Manta (Chen et al., 2016) as described in (Demidov et al., 2024). SNVs, indels, SVs, and CNVs in long-read genome sequencing data were detected and filtered as described in (Steyaert et al., 2025). Chromosomal aberrations were detected by optical genome mapping and prioritized as described in (Mantere et al., 2021). SNVs and indels were assigned to a gene if they were within the gene body ± 5 Kbp upstream and downstream, as suggested by VEP (Zerbino et al., 2018). SVs and chromosomal aberration calls were assigned to a gene if they had any overlap with the gene body. Repeat expansions were called using ExpansionHunter (Dolzhenko et al., 2017) and prioritized as described in (Sanden et al., 2021).

### Phenotype scores

First, the Human Phenotype Ontology (HPO) file was downloaded from The HPO Project portal (hpo.jax.org) from the June 2024 Release (Köhler et al., 2020). Then, the semantic similarity score was computed between all available HPO terms on that release and the HPO terms from each individual using the Resnik method, which returns the information content of the most informative common ancestor (using the compareHPSets function from the R package Phenotype Consensus Analysis PCAN (Godard and Page, 2016)). These scores were grouped by gene using the phenotype-to-genes matrix downloaded from the same portal. Finally, a single aggregate semantic similarity score was computed per gene-individual combination using the average of the best phenotype matches (through the hpSetCompSummary function from the R package PCAN).

### Results integration

The RNA-seq results were integrated with the genetic variants and phenotype scores into one table per family for which each row is a gene-sample combination. The combined results table consisted of all RNA outliers per sample including expression fold change, splicing differential; the two most impactful (according to VEP) rare SNVs and indels; overlap with a rare CNV, SV, repeat expansion; phenotype semantic similarity scores, and annotation of OMIM and ERN disease genes. To reduce noise in samples with more than 20 expression or splicing outliers, we prioritized genes with a phenotype score above 3 or those carrying at least one rare variant. A per-family report was generated containing the filtered results tables and the most informative visualizations. For expression, the figures included z-score versus genomic position and fold change distributions. For splicing, split-read support versus total split reads and Sashimi plots were produced to illustrate exon usage and junction-level alterations.

### Classification framework

The RNA outliers classification framework is based on four levels: RNA evidence, gene-phenotype association, variant mechanism, and segregation. Different combinations of these levels indicate the final outlier classification as a very strong diagnostic candidate, diagnostic candidate, new disease gene candidate, functionally unclear gene, or discarded. The interpreter could select each of the four categories in a drop-down menu implemented in a spreadsheet, and the final classification was automatically computed.

#### RNA evidence

This category has two values: reliable or unreliable. Besides the statistical significance (based on adjusted p-value and differential splicing), visualizing the results and aligned data was required to categorize each finding. For expression, volcano plots, Manhattan plots, and fold change plots (Fig. 6) were assessed. For splicing, Sashimi plots (Fig. 7) and the fitted delta Jaccard index were evaluated. All the figures were generated using OUTRIDER or FRASER. Sashimi plots were additionally created using the Integrative Genomics Viewer (IGV) as it offers more flexibility than FRASER. Reliable expression or splicing aberrations match the ACMG criterion pathogenic strong (PS) 3: “well-established functional studies show a deleterious effect” (Richards et al., 2015), in agreement with (Bournazos et al., 2022).

Unreliable aberrant splicing events are frequently present in regions showing excessive splicing activity or long-range splicing patterns, which may reflect repetitive sequences or high sequence homology, such as gene-pseudogene pairs, gene families, or transposable elements.

#### Gene-phenotype association

This assessment was classified into the following categories: “disease gene with a matching phenotype”, “disease gene with a compatible phenotype”, “non-disease gene, but with a function consistent with the phenotype”, “disease gene with unclear phenotype association”, and “unrelated”. Disease genes were defined as genes with a documented disease association in OMIM (Amberger et al., 2019). A matching phenotype was defined in accordance with the ACMG criterion PP4 (pathogenic supporting evidence based on phenotype, (Richards et al., 2015)). We additionally included the category “compatible phenotype” to capture cases of this research study in which the clinical presentation of the patient did not fully overlap with the currently described phenotypic spectrum of the gene.

The category “disease gene with unclear phenotype association” was introduced to include genes for which, based on currently known phenotypic associations, it was not possible to determine whether they were concordant with the patient’s clinical presentation. The “unrelated” category included both disease-associated and non–disease-associated genes whose known functions or reported phenotypes showed no apparent relationship to the patient’s observed phenotype.

#### Variant mechanism

This level of evidence comprises four categories: consistent variant mechanism with the RNA outlier, unclear variant mechanism, no rare variant found for this gene (see subsection Variant Annotation), or discarded after experimental assessment. In addition to the evidence for variant pathogenicity described in Table 3 of the ACMG guidelines (Richards et al., 2015), we evaluated the consistency between plausible mechanistic effects of the genetic variant and the type of RNA-seq aberrant event. Variants expected to trigger nonsense-mediated decay, including stop-gain, frameshift, or canonical splice-site variants, as well as splicing defects that introduce a frameshift or a premature termination codon, were considered consistent with reduced gene expression. Copy number gains were considered consistent with increased expression, whereas deletions were consistent with reduced expression. Variants located near splice sites exhibiting any kind of splicing aberration, or within deep intronic regions nearby novel or elongated exons, as well as exonic variants proximal to truncating events, were also regarded as mechanistically consistent. This is in line with the ClinGen SVI Splicing Subgroup recommendations (Walker et al., 2023) that suggest to assign those variants to a new category called PVS1_Strength (RNA) and with the SpliceACORD recommendations (Bournazos et al., 2022), which assign these variants to PS3. In addition, deletions spanning multiple exons were considered capable of inducing exon truncation or exon skipping.

#### Segregation

Five segregation classes were defined: “consistent”, “inconsistent”, “inconclusive”, and “not available”. We considered segregation to be “consistent” if the variants co-segregated in multiple affected family members (Richards et al., 2015). For families with RNA-seq data for multiple family members, we further required that the observed expression or splicing defect co-segregated with the phenotype (same type of event, same direction, nominal significance for other cases). One recent example of segregated exon creation is described in (van der Westhuizen et al., 2025).

## Data Availability

Raw data will be available at the European Genome-Phenome Archive (https://ega-archive.org/datasets/) under the Solve-RD study EGAS00001003851, and can be accessed following approval from the Solve-RD Data Access Committee.

## Data & code availability

Raw data will be available at the European Genome-Phenome Archive (https://ega-archive.org/datasets/) under the Solve-RD study EGAS00001003851, and can be accessed following approval from the Solve-RD Data Access Committee. The patient consent of the muscle samples from IDIBAPS and the PBMC samples from IDIBELL do not allow sharing of data.

## Ethics declaration

The ethics committee of University Hospital of Tübingen gave ethical approval for this work (ClinicalTrials.gov ID: NCT03491280, https://clinicaltrials.gov/study/NCT03491280). Informed consent for data sharing, including indirect identifiers within Europe for research, was obtained from all recruited individuals. All data submitters confirmed the code of conduct of RD-connect GPAP. This study adheres to the principles set out in the Declaration of Helsinki.

## Acknowledgements

We are indebted to the affected individuals and their families. We would like to thank Marlen C. Lauffer for advice on the possibility of ASO treatment for the solved cases.

The Solve-RD project has received funding from the European Union’s Horizon 2020 research and innovation programme under grant agreement No 779257. Solve-RD research is supported (not financially) by ERN ITHACA (project ID no. 101085231), ERN RND (project ID no. 101155994), ERN EURO-NMD (project ID no. 101156434), ERN EpiCARE (project ID 101156811), and ERN RITA (project ID 101155878). All ERNs are cofunded by the European Union within the framework of the Third Health Programme. ERDERA has received funding from the European Union’s Horizon Europe research and innovation programme under grant agreement N°101156595. Views and opinions expressed are those of the author(s) only and do not necessarily reflect those of the European Union or any other granting authority, who cannot be held responsible for them.

VAY, RL, CM and JG were supported by the Deutsche Forschungsgemeinschaft (German Research Foundation) via the project NFDI 1/1 ‘GHGA — German Human Genome–Phenome Archive’ (441914366). The TUM IT infrastructure was cofunded via the Deutsche Forschungsgemeinschaft (German Research Foundation, project ID 461264291). BEA was supported by the predoctoral program “Joan Oró” of the Secretary of Universities and Research of the Department of Research and Universities of the Government of Catalonia with codes 2024 FI-1 00075 and 2025 FI-2 00075, co-financed by the European Union. HM was supported by the Wellcome Trust grant 220906/Z/20/Z and UCL Global Engagement Fund scheme (2022/23 GEF project). HL receives support from the Canadian Institutes of Health Research (CIHR) for Foundation Grant FDN-167281 (Precision Health for Neuromuscular Diseases), Transnational Team Grant ERT-174211 (ProDGNE) and Network Grant OR2-189333 (NMD4C), from the Canada Foundation for Innovation (CFI-JELF 38412), the Canada Research Chairs program (Canada Research Chair in Neuromuscular Genomics and Health, 950-232279), the European Commission (Grant # 101080249) and the Canada Research Coordinating Committee New Frontiers in Research Fund (NFRFG-2022-00033) for SIMPATHIC, and from the Government of Canada First Research Excellence Fund (CFREF) for the Brain-Heart Interconnectome (CFREF-2022-00007). KP is a recipient of a Canadian Institutes of Health Research (CIHR) postdoctoral fellowship award under award no: MFE-491707. JP was supported by the Else-Kröner-Fresenius-Stiftung Clinician Scientist program “precise.net” and by the intramural TÜFF program (3049-0-0). AP has received funding from the Secretariat for Universities and Research of the Ministry of Business and Knowledge of the Government of Catalonia (2021SGR00899), and the Instituto de Salud Carlos III (ISCIII) (FIS PI23/00835) ‘Fondo Europeo de Desarrollo Regional (FEDER), Unión Europea, una manera de hacer Europa’. ASC was supported by the grants FPU20/06692 and EST23/00463 from “Ministerio de Universidades” (Spain). AH was supported by a ZonMW (The Netherlands Organization for Health Research and Development) Vici grant (No. 09150182310053). KL receives support from the German Research Foundation (DFG, LO1555/10-1). DNdB was supported by Instituto de Salud Carlos III (Grant CP22/00141).

Institutional support to CNAG was provided by the Spanish Ministry of Science and Innovation through the Instituto de Salud Carlos III, and by the Generalitat de Catalunya through the Departament de Salut and the Departament de Recerca i Universitats. We thank the UMCG Genomics Coordination Center, the MOLGENIS team, the University of Groningen Center for Information Technology, the European Bioinformatics Institute and the UMCG research IT program and their sponsors for providing data infrastructure and support.

## References

Aicher, J. K., Jewell, P., Vaquero-Garcia, J., Barash, Y., and Bhoj, E. J. (2020). Mapping RNA splicing variations in clinically accessible and nonaccessible tissues to facilitate Mendelian disease diagnosis using RNA-seq. Genet. Med. 22, 1181–1190. doi: 10.1038/s41436-020-0780-y

Amberger, J. S., Bocchini, C. A., Scott, A. F., and Hamosh, A. (2019). OMIM.org: leveraging knowledge across phenotype–gene relationships. Nucleic Acids Res. 47, D1038–D1043. doi: 10.1093/nar/gky1151

Andersen, E. F., Azzariti, D. R., Babb, L., Berg, J. S., Biesecker, L. G., Bly, Z., et al. (2025). The Clinical Genome Resource (ClinGen): Advancing genomic knowledge through global curation. Genet. Med. 27. doi: 10.1016/j.gim.2024.101228

Arriaga, T. M., Mendez, R., Ungar, R. A., Bonner, D. E., Matalon, D. R., Lemire, G., et al. (2025). Transcriptome-wide outlier approach identifies individuals with minor spliceopathies. Am. J. Hum. Genet. 112, 2458–2475. doi: 10.1016/j.ajhg.2025.08.018

Bertoli-Avella, A. M., Radefeldt, M., Al-Ali, R., Pardo, L. M., Lemke, S., Leubauer, A., et al. (2025). Beyond genomics: using RNA-seq from dried blood spots to unlock the clinical relevance of splicing variation in a diagnostic setting. Eur. J. Hum. Genet. 33, 614–623. doi: 10.1038/s41431-025-01792-2

Bournazos, A. M., Riley, L. G., Bommireddipalli, S., Ades, L., Akesson, L. S., Al-Shinnag, M., et al. (2022). Standardized practices for RNA diagnostics using clinically accessible specimens reclassifies 75% of putative splicing variants. Genet. Med. 24, 130–145. doi: 10.1016/j.gim.2021.09.001

Boycott, K. M., Hartley, T., Kernohan, K. D., Dyment, D. A., Howley, H., Innes, A. M., et al. (2022). Care4Rare Canada: Outcomes from a decade of network science for rare disease gene discovery. Am. J. Hum. Genet. 109, 1947–1959. doi: 10.1016/j.ajhg.2022.10.002

Brechtmann, F., Mertes, C., Matusevičiūtė, A., Yépez, V. A., Avsec, Ž., Herzog, M., et al. (2018). OUTRIDER: A Statistical Method for Detecting Aberrantly Expressed Genes in RNA Sequencing Data. Am. J. Hum. Genet. 103, 907–917. doi: 10.1016/j.ajhg.2018.10.025

Cheerie, D., Meserve, M. M., Beijer, D., Kaiwar, C., Newton, L., Tavares, A. L. T., et al. (2025). Consensus guidelines for assessing eligibility of pathogenic DNA variants for antisense oligonucleotide treatments. Am. J. Hum. Genet. 112, 975–983. doi: 10.1016/j.ajhg.2025.02.017

Chen, X., Schulz-Trieglaff, O., Shaw, R., Barnes, B., Schlesinger, F., Källberg, M., et al. (2016). Manta: rapid detection of structural variants and indels for germline and cancer sequencing applications. Bioinformatics 32, 1220–1222. doi: 10.1093/bioinformatics/btv710

Chen, Y., Dawes, R., Kim, H. C., Ljungdahl, A., Stenton, S. L., Walker, S., et al. (2024). De novo variants in the RNU4-2 snRNA cause a frequent neurodevelopmental syndrome. Nature 632, 832–840. doi: 10.1038/s41586-024-07773-7

Chui, M. M.-C., Kwong, A. K.-Y., Leung, H. Y. C., Pang, C., Scheller, I. F., Wong, S. S.-N., et al. (2025). An outlier approach: advancing diagnosis of neurological diseases through integrating proteomics into multi-omics guided exome reanalysis. Npj Genomic Med. 10, 36. doi: 10.1038/s41525-025-00493-5

Cohen, E., Bonne, G., Rivier, F., and Hamroun, D. (2021). The 2022 version of the gene table of neuromuscular disorders (nuclear genome). Neuromuscul. Disord. 31, 1313–1357. doi: 10.1016/j.nmd.2021.11.004

Cordts, I., Semmler, L., Prasuhn, J., Seibt, A., Herebian, D., Navaratnarajah, T., et al. (2022). Bi-Allelic COQ4 Variants Cause Adult-Onset Ataxia-Spasticity Spectrum Disease. Mov. Disord. 37, 2147–2153. doi: 10.1002/mds.29167

Cummings, B. B., Marshall, J. L., Tukiainen, T., Lek, M., Donkervoort, S., Foley, A. R., et al. (2017). Improving genetic diagnosis in Mendelian disease with transcriptome sequencing. Sci. Transl. Med. 9, eaal5209. doi: 10.1126/scitranslmed.aal5209

Dawood, M., Heavner, B., Wheeler, M. M., Ungar, R. A., LoTempio, J., Wiel, L., et al. (2025). GREGoR: accelerating genomics for rare diseases. Nature 647, 331–342. doi: 10.1038/s41586-025-09613-8

De Cock, L., D’haenens, E., Vantomme, L., Backers, L., Beyens, A., Claes, K. B., et al. (2025). Cracking rare disorders: a new minimally invasive RNA-seq protocol. Npj Genomic Med. 10, 45. doi: 10.1038/s41525-025-00502-7

Demidov, G., Laurie, S., Torella, A., Piluso, G., Scala, M., Morleo, M., et al. (2024). Structural variant calling and clinical interpretation in 6224 unsolved rare disease exomes. Eur. J. Hum. Genet. 32, 998–1004. doi: 10.1038/s41431-024-01637-4

Deshwar, A. R., Yuki, K. E., Hou, H., Liang, Y., Khan, T., Celik, A., et al. (2023). Trio RNA sequencing in a cohort of medically complex children. Am. J. Hum. Genet. 110, 895–900. doi: 10.1016/j.ajhg.2023.03.006

Dobin, A., Davis, C. A., Schlesinger, F., Drenkow, J., Zaleski, C., Jha, S., et al. (2013). STAR: ultrafast universal RNA-seq aligner. Bioinformatics 29, 15–21. doi: 10.1093/bioinformatics/bts635

Dolzhenko, E., van Vugt, J. J. F. A., Shaw, R. J., Bekritsky, M. A., van Blitterswijk, M., Narzisi, G., et al. (2017). Detection of long repeat expansions from PCR-free whole-genome sequence data. Genome Res. 27, 1895–1903. doi: 10.1101/gr.225672.117

Estévez-Arias, B., Matalonga, L., Yubero, D., Polavarapu, K., Codina, A., Ortez, C., et al. (2024). Phenotype-driven genomics enhance diagnosis in children with unresolved neuromuscular diseases. Eur. J. Hum. Genet. 33, 239–247. doi: 10.1038/s41431-024-01699-4

Fazal, S., Danzi, M. C., van Kuilenburg, A. B. P., Reich, S., Traschütz, A., Bender, B., et al. (2023). Repeat expansions nested within tandem CNVs: a unique structural change in GLS exemplifies the diagnostic challenges of non-coding pathogenic variation. Hum. Mol. Genet. 32, 46–54. doi: 10.1093/hmg/ddac173

Frankish, A., Diekhans, M., Jungreis, I., Lagarde, J., Loveland, J. E., Mudge, J. M., et al. (2021). GENCODE 2021. Nucleic Acids Res. 49, D916–D923. doi: 10.1093/nar/gkaa1087

Frésard, L., Smail, C., Ferraro, N. M., Teran, N. A., Li, X., Smith, K. S., et al. (2019). Identification of rare-disease genes using blood transcriptome sequencing and large control cohorts. Nat. Med. 25, 911–919. doi: 10.1038/s41591-019-0457-8

García-Alcalde, F., Okonechnikov, K., Carbonell, J., Cruz, L. M., Götz, S., Tarazona, S., et al. (2012). Qualimap: evaluating next-generation sequencing alignment data. Bioinformatics 28, 2678–2679. doi: 10.1093/bioinformatics/bts503

Godard, P., and Page, M. (2016). PCAN: phenotype consensus analysis to support disease-gene association. BMC Bioinformatics 17, 518. doi: 10.1186/s12859-016-1401-2

Gonorazky, H. D., Naumenko, S., Ramani, A. K., Nelakuditi, V., Mashouri, P., Wang, P., et al. (2019). Expanding the Boundaries of RNA Sequencing as a Diagnostic Tool for Rare Mendelian Disease. Am. J. Hum. Genet. 104, 466–483. doi: 10.1016/j.ajhg.2019.01.012

Groh, M., Lufino, M. M. P., Wade-Martins, R., and Gromak, N. (2014). R-loops Associated with Triplet Repeat Expansions Promote Gene Silencing in Friedreich Ataxia and Fragile X Syndrome. PLOS Genet. 10, e1004318. doi: 10.1371/journal.pgen.1004318

Hart, R. K., Fokkema, I. F. A. C., DiStefano, M., Hastings, R., Laros, J. F. J., Taylor, R., et al. (2024). HGVS Nomenclature 2024: improvements to community engagement, usability, and computability. Genome Med. 16, 149. doi: 10.1186/s13073-024-01421-5

HGVS Variant Nomenclature Committee (2025). Splicing - HGVS Nomenclature. Splicing. Available at: https://hgvs-nomenclature.org/stable/recommendations/RNA/splicing/ (Accessed December 1, 2025).

Hong, S. E., Kneissl, J., Cho, A., Kim, M. J., Park, S., Lee, J., et al. (2022). Transcriptome-based variant calling and aberrant mRNA discovery enhance diagnostic efficiency for neuromuscular diseases. J. Med. Genet. 59, 1075–1081. doi: 10.1136/jmedgenet-2021-108307

Höps, W., Weiss, M. M., Derks, R., Galbany, J. C., Ouden, A. den, Heuvel, S. van den, et al. (2025). HiFi long-read genomes for difficult-to-detect, clinically relevant variants. Am. J. Hum. Genet. 112, 450–456. doi: 10.1016/j.ajhg.2024.12.013

Jackson, A., Blakes, A. J., Wall, E., Clarke, N., Abdelhadi, O., Agrawal, S., et al. (2025). Biallelic variants in RNU2-2 cause a remarkably frequent developmental epileptic encephalopathy. 2025.09.02.25334957. doi: 10.1101/2025.09.02.25334957

Jaramillo Oquendo, C., Wai, H. A., Rich, W. I., Bunyan, D. J., Thomas, N. S., Hunt, D., et al. (2024). Identification of diagnostic candidates in Mendelian disorders using an RNA sequencing-centric approach. Genome Med. 16, 110. doi: 10.1186/s13073-024-01381-w

Jensen, T. D., Ni, B., Reuter, C. M., Gorzynski, J. E., Fazal, S., Bonner, D., et al. (2025). Integration of transcriptomics and long-read genomics prioritizes structural variants in rare disease. Genome Res. 35, 914–928. doi: 10.1101/gr.279323.124

Kernohan, K. D., and Boycott, K. M. (2024). The expanding diagnostic toolbox for rare genetic diseases. Nat. Rev. Genet. 25, 401–415. doi: 10.1038/s41576-023-00683-w

Kim, J., Hu, C., Moufawad El Achkar, C., Black, L. E., Douville, J., Larson, A., et al. (2019). Patient-Customized Oligonucleotide Therapy for a Rare Genetic Disease. N. Engl. J. Med. 381, 1644–1652. doi: 10.1056/NEJMoa1813279

Kim, J.-H., Shinde, D. N., Reijnders, M. R. F., Hauser, N. S., Belmonte, R. L., Wilson, G. R., et al. (2016). De Novo Mutations in SON Disrupt RNA Splicing of Genes Essential for Brain Development and Metabolism, Causing an Intellectual-Disability Syndrome. Am. J. Hum. Genet. 99, 711–719. doi: 10.1016/j.ajhg.2016.06.029

Kobayashi, H., Abe, K., Matsuura, T., Ikeda, Y., Hitomi, T., Akechi, Y., et al. (2011). Expansion of intronic GGCCTG hexanucleotide repeat in NOP56 causes SCA36, a type of spinocerebellar ataxia accompanied by motor neuron involvement. Am. J. Hum. Genet. 89, 121–130. doi: 10.1016/j.ajhg.2011.05.015

Köhler, S., Gargano, M., Matentzoglu, N., Carmody, L. C., Lewis-Smith, D., Vasilevsky, N. A., et al. (2020). The Human Phenotype Ontology in 2021. Nucleic Acids Res. 49, D1207–D1217. doi: 10.1093/nar/gkaa1043

Kremer, L. S., Bader, D. M., Mertes, C., Kopajtich, R., Pichler, G., Iuso, A., et al. (2017). Genetic diagnosis of Mendelian disorders via RNA sequencing. Nat. Commun. 8, 15824. doi: 10.1038/ncomms15824

Krueger, F. (2021). TrimGalore: A wrapper around Cutadapt and FastQC to consistently apply adapter and quality trimming to FastQ files, with extra functionality for RRBS data. Available at: https://github.com/FelixKrueger/TrimGalore?tab=readme-ov-file (Accessed October 3, 2024).

Kumar, R., Corbett, M. A., Smith, N. J. C., Hock, D. H., Kikhtyak, Z., Semcesen, L. N., et al. (2022). Oligonucleotide correction of an intronic TIMMDC1 variant in cells of patients with severe neurodegenerative disorder. Npj Genomic Med. 7, 1–12. doi: 10.1038/s41525-021-00277-7

Laurie, S., Steyaert, W., de Boer, E., Polavarapu, K., Schuermans, N., Sommer, A. K., et al. (2025). Genomic reanalysis of a pan-European rare-disease resource yields new diagnoses. Nat. Med. 31, 478–489. doi: 10.1038/s41591-024-03420-w

Lawrence, M., Huber, W., Pagès, H., Aboyoun, P., Carlson, M., Gentleman, R., et al. (2013). Software for Computing and Annotating Genomic Ranges. PLoS Comput. Biol. 9, e1003118. doi: 10.1371/journal.pcbi.1003118

Lee, M., Kwong, A. K. Y., Chui, M. M. C., Chau, J. F. T., Mak, C. C. Y., Au, S. L. K., et al. (2022). Diagnostic potential of the amniotic fluid cells transcriptome in deciphering mendelian disease: a proof-of-concept. Npj Genomic Med. 7, 74. doi: 10.1038/s41525-022-00347-4

Li, S., Zhao, S., Sinson, J. C., Bajic, A., Rosenfeld, J. A., Neeley, M. B., et al. (2024). The clinical utility and diagnostic implementation of human subject cell transdifferentiation followed by RNA sequencing. Am. J. Hum. Genet. 111, 841–862. doi: 10.1016/j.ajhg.2024.03.007

Liao, Y., Smyth, G. K., and Shi, W. (2019). The R package Rsubread is easier, faster, cheaper and better for alignment and quantification of RNA sequencing reads. Nucleic Acids Res. 47, e47. doi: 10.1093/nar/gkz114

Lindstrand, A., Lagerstedt-Robinson, K., Jemt, A., Kvarnung, M., Ygberg, S., Vonlanthen, S., et al. (2025). The Genomic Medicine Center Karolinska 10-year report on genome sequencing for rare diseases and a strategy for stepwise clinical implementation. doi: 10.21203/rs.3.rs-6790162/v1

Liu, Y. H., Luo, C., Golding, S. G., Ioffe, J. B., and Zhou, X. M. (2024). Tradeoffs in alignment and assembly-based methods for structural variant detection with long-read sequencing data. Nat. Commun. 15, 2447. doi: 10.1038/s41467-024-46614-z

Lunke, S., Bouffler, S. E., Patel, C. V., Sandaradura, S. A., Wilson, M., Pinner, J., et al. (2023). Integrated multi-omics for rapid rare disease diagnosis on a national scale. Nat. Med. 29, 1681–1691. doi: 10.1038/s41591-023-02401-9

Luo, X., Xiao, B., Liang, L., Zhang, K., Xu, T., Liu, H., et al. (2025). Blood RNA-seq in rare disease diagnostics: a comparative study of cases with and without candidate variants. J. Transl. Med. 23, 586. doi: 10.1186/s12967-025-06609-w

Malik, I., Kelley, C. P., Wang, E. T., and Todd, P. K. (2021). Molecular mechanisms underlying nucleotide repeat expansion disorders. Nat. Rev. Mol. Cell Biol. 22, 589–607. doi: 10.1038/s41580-021-00382-6

Mantere, T., Neveling, K., Pebrel-Richard, C., Benoist, M., Zande, G. van der, Kater-Baats, E., et al. (2021). Optical genome mapping enables constitutional chromosomal aberration detection. Am. J. Hum. Genet. 108, 1409–1422. doi: 10.1016/j.ajhg.2021.05.012

Marchant, R. G., Bryen, S. J., Bahlo, M., Cairns, A., Chao, K. R., Corbett, A., et al. (2024). Genome and RNA sequencing boost neuromuscular diagnoses to 62% from 34% with exome sequencing alone. Ann. Clin. Transl. Neurol. 11, 1250–1266. doi: 10.1002/acn3.52041

Martin-Geary, A. C., Blakes, A. J. M., Dawes, R., Findlay, S. D., Lord, J., Dong, S., et al. (2025). Systematic identification of disease-causing promoter and untranslated region variants in 8040 undiagnosed individuals with rare disease. Genome Med. 17, 40. doi: 10.1186/s13073-025-01464-2

Marwaha, S., Knowles, J. W., and Ashley, E. A. (2022). A guide for the diagnosis of rare and undiagnosed disease: beyond the exome. Genome Med. 14, 23. doi: 10.1186/s13073-022-01026-w

Miller, D. E., Sulovari, A., Wang, T., Loucks, H., Hoekzema, K., Munson, K. M., et al. (2021). Targeted long-read sequencing identifies missing disease-causing variation. Am. J. Hum. Genet. 108, 1436–1449. doi: 10.1016/j.ajhg.2021.06.006

Montgomery, S. B., Bernstein, J. A., and Wheeler, M. T. (2022). Toward transcriptomics as a primary tool for rare disease investigation. Mol. Case Stud. 8, a006198. doi: 10.1101/mcs.a006198

Morales, J., Pujar, S., Loveland, J. E., Astashyn, A., Bennett, R., Berry, A., et al. (2022). A joint NCBI and EMBL-EBI transcript set for clinical genomics and research. Nature 604, 310–315. doi: 10.1038/s41586-022-04558-8

Müller, J. S., Rabinowicz, S., Zaharieva, I., Pini, V., Yépez, V. A., Esteve-Codina, A., et al. (2025). Multi-omics approach identifies a novel recessive pathogenic variant in the TNNT3 gene in two siblings with congenital myopathy. Neuromuscul. Disord. 52, 105415. doi: 10.1016/j.nmd.2025.105415

Murdock, D. R., Dai, H., Burrage, L. C., Rosenfeld, J. A., Ketkar, S., Müller, M. F., et al. (2021). Transcriptome-directed analysis for Mendelian disease diagnosis overcomes limitations of conventional genomic testing. J. Clin. Invest. 131, e141500. doi: 10.1172/JCI141500

Nava, C., Cogne, B., Santini, A., Leitão, E., Lecoquierre, F., Chen, Y., et al. (2025). Dominant variants in major spliceosome U4 and U5 small nuclear RNA genes cause neurodevelopmental disorders through splicing disruption. Nat. Genet. 57, 1374–1388. doi: 10.1038/s41588-025-02184-4

Neuhoff, K., Kilicarslan, O. A., Preuße, C., Zaum, A.-K., Kölbel, H., Lochmüller, H., et al. (2024). Expanding the Molecular Genetic Landscape of Dystrophinopathies and Associated Phenotypes. Biomedicines 12, 2738. doi: 10.3390/biomedicines12122738

Nicolas-Martinez, E. C., Robinson, O., Pflueger, C., Gardner, A., Corbett, M. A., Ritchie, T., et al. (2024). RNA variant assessment using transactivation and transdifferentiation. Am. J. Hum. Genet. 111, 1673–1699. doi: 10.1016/j.ajhg.2024.06.018

Pogue, R. E., Cavalcanti, D. P., Shanker, S., Andrade, R. V., Aguiar, L. R., de Carvalho, J. L., et al. (2018). Rare genetic diseases: update on diagnosis, treatment and online resources. Drug Discov. Today 23, 187–195. doi: 10.1016/j.drudis.2017.11.002

Richards, S., Aziz, N., Bale, S., Bick, D., Das, S., Gastier-Foster, J., et al. (2015). Standards and guidelines for the interpretation of sequence variants: a joint consensus recommendation of the American College of Medical Genetics and Genomics and the Association for Molecular Pathology. Genet. Med. 17, 405–423. doi: 10.1038/gim.2015.30

Riquin, K., Isidor, B., Mercier, S., Nizon, M., Colin, E., Bonneau, D., et al. (2024). Integrating RNA-Seq into genome sequencing workflow enhances the analysis of structural variants causing neurodevelopmental disorders. J. Med. Genet. 61, 47–56. doi: 10.1136/jmg-2023-109263

Roberts, T. C., Langer, R., and Wood, M. J. A. (2020). Advances in oligonucleotide drug delivery. Nat. Rev. Drug Discov. 19, 673–694. doi: 10.1038/s41573-020-0075-7

Sanden, B. P. G. H. van der, Corominas, J., Groot, M. de, Pennings, M., Meijer, R. P. P., Verbeek, N., et al. (2021). Systematic analysis of short tandem repeats in 38,095 exomes provides an additional diagnostic yield. Genet. Med. 23, 1569–1573. doi: 10.1038/s41436-021-01174-1

Saparov, A., Dzinovic, I., Brunet, T., Yépez, V. A., Hölzlwimmer, F., Indelicato, E., et al. (2026). Fibroblast Transcriptomics in Molecular Diagnostics of a Comprehensive Dystonia Cohort. Ann. Neurol. doi: 10.1002/ana.78171

Scheller, I. F., Lutz, K., Mertes, C., Yépez, V. A., and Gagneur, J. (2023). Improved detection of aberrant splicing with FRASER 2.0 and the intron Jaccard index. Am. J. Hum. Genet. 110, 2056–2067. doi: 10.1016/j.ajhg.2023.10.014

Segarra-Casas, A., Domínguez-González, C., Natera-de Benito, D., Kapetanovic, S., Hernández-Laín, A., Estévez-Arias, B., et al. (2025). Translating Muscle RNAseq Into the Clinic for the Diagnosis of Muscle Diseases. Ann. Clin. Transl. Neurol. 12, 1465–1479. doi: 10.1002/acn3.70078

Segarra-Casas, A., Yépez, V. A., Demidov, G., Laurie, S., Esteve-Codina, A., Gagneur, J., et al. (2024). An Integrated Transcriptomics and Genomics Approach Detects an X/Autosome Translocation in a Female with Duchenne Muscular Dystrophy. Int. J. Mol. Sci. 25, 7793. doi: 10.3390/ijms25147793

Smirnov, D., Schlieben, L. D., Peymani, F., Berutti, R., and Prokisch, H. (2022). Guidelines for clinical interpretation of variant pathogenicity using RNA phenotypes. Hum. Mutat. 43, 1056–1070. doi: 10.1002/humu.24416

Sommer, A. K., te Paske, I. B. A. W., Jansen, E. A. M., Gschwind, A., Demidov, G., Steinke-Lange, V., et al. (2026). Mutational Landscape of Colorectal Tumors From Individuals With Unexplained Adenomatous or Serrated Colorectal Polyposis. Gastroenterology. doi: 10.1053/j.gastro.2025.10.011

Stark, J. C., Pipko, N., Liang, Y., Szuto, A., Tsoi, C. T., Dickson, M. A., et al. (2025). Clinical applications of and molecular insights from RNA sequencing in a rare disease cohort. Genome Med. 17, 72. doi: 10.1186/s13073-025-01494-w

Stark, Z., Boughtwood, T., Haas, M., Braithwaite, J., Gaff, C. L., Goranitis, I., et al. (2023). Australian Genomics: Outcomes of a 5-year national program to accelerate the integration of genomics in healthcare. Am. J. Hum. Genet. 110, 419–426. doi: 10.1016/j.ajhg.2023.01.018

Steyaert, W., Sagath, L., Demidov, G., Yépez, V. A., Esteve-Codina, A., Gagneur, J., et al. (2025). Unraveling undiagnosed rare disease cases by HiFi long-read genome sequencing. Genome Res. 35, 755–768. doi: 10.1101/gr.279414.124

Tavtigian, S. V., Greenblatt, M. S., Harrison, S. M., Nussbaum, R. L., Prabhu, S. A., Boucher, K. M., et al. (2018). Modeling the ACMG/AMP variant classification guidelines as a Bayesian classification framework. Genet. Med. 20, 1054–1060. doi: 10.1038/gim.2017.210

Tesi, B., Boileau, C., Boycott, K. M., Canaud, G., Caulfield, M., Choukair, D., et al. (2023). Precision medicine in rare diseases: What is next? J. Intern. Med. 294, 397–412. doi: 10.1111/joim.13655

The 100,000 Genomes Project Pilot Investigators (2021). 100,000 Genomes Pilot on Rare-Disease Diagnosis in Health Care — Preliminary Report. N. Engl. J. Med. 385, 1868–1880. doi: 10.1056/NEJMoa2035790

Tumiene, B., Graessner, H., Mathijssen, I. M., Pereira, A. M., Schaefer, F., Scarpa, M., et al. (2021). European Reference Networks: challenges and opportunities. J. Community Genet. 12, 217–229. doi: 10.1007/s12687-021-00521-8

van der Westhuizen, J., Yépez, V. A., and Moosa, S. (2025). RNA Sequencing for Rare Disease Diagnosis in a South African Family: A Novel Exon Elongation Event in OFD1. Am. J. Med. Genet. A. n/a. doi: 10.1002/ajmga.70024

Vialle, R. A., de Paiva Lopes, K., Bennett, D. A., Crary, J. F., and Raj, T. (2022). Integrating whole-genome sequencing with multi-omic data reveals the impact of structural variants on gene regulation in the human brain. Nat. Neurosci. 25, 504–514. doi: 10.1038/s41593-022-01031-7

Walker, L. C., Hoya, M. de la, Wiggins, G. A. R., Lindy, A., Vincent, L. M., Parsons, M. T., et al. (2023). Using the ACMG/AMP framework to capture evidence related to predicted and observed impact on splicing: Recommendations from the ClinGen SVI Splicing Subgroup. Am. J. Hum. Genet. 110, 1046–1067. doi: 10.1016/j.ajhg.2023.06.002

Wojcik, M., Lemire, G., Berger, E., Zaki Maha S., Wissmann Mariel, Win Wathone, et al. (2024). Genome Sequencing for Diagnosing Rare Diseases. N. Engl. J. Med. 390, 1985–1997. doi: 10.1056/NEJMoa2314761

Xiao, B., Luo, X., Liu, Y., Ye, H., Liu, H., Fan, Y., et al. (2024). Combining optical genome mapping and RNA-seq for structural variants detection and interpretation in unsolved neurodevelopmental disorders. Genome Med. 16, 113. doi: 10.1186/s13073-024-01382-9

Yang, Y., Muzny, D. M., Reid, J. G., Bainbridge, M. N., Willis, A., Ward, P. A., et al. (2013). Clinical Whole-Exome Sequencing for the Diagnosis of Mendelian Disorders. N. Engl. J. Med. 369, 1502–1511. doi: 10.1056/NEJMoa1306555

Yépez, V. A., Demidov, G., Ellwanger, K., Laurie, S., Luknárová, R., Joseph Maran, M. I., et al. (2025). The Solve-RD Solvathons as a pan-European interdisciplinary collaboration to diagnose patients with rare disease. Nat. Genet. 59, 2361–2370. doi: 10.1038/s41588-025-02290-3

Yépez, V. A., Gusic, M., Kopajtich, R., Mertes, C., Smith, N. H., Alston, C. L., et al. (2022). Clinical implementation of RNA sequencing for Mendelian disease diagnostics. Genome Med. 14, 38. doi: 10.1186/s13073-022-01019-9

Yépez, V. A., Mertes, C., Müller, M. F., Klaproth-Andrade, D., Wachutka, L., Frésard, L., et al. (2021). Detection of aberrant gene expression events in RNA sequencing data. Nat. Protoc. 16, 1276–1296. doi: 10.1038/s41596-020-00462-5

Yepez, V., Gusic, M., Kopajtich, R., Meitinger, T., Prokisch, H., and Gagneur, J. (2022). Gene expression and splicing counts from the Yepez, Gusic et al study - fibroblast, hg19, strand-specific, high seq depth. doi: 10.5281/zenodo.7510836

Youssefian, L., Saeidian, A. H., Palizban, F., Bagherieh, A., Abdollahimajd, F., Sotoudeh, S., et al. (2021). Whole-Transcriptome Analysis by RNA Sequencing for Genetic Diagnosis of Mendelian Skin Disorders in the Context of Consanguinity. Clin. Chem. 67, 876–888. doi: 10.1093/clinchem/hvab042

Zardetto, B., Lauffer, M. C., van Roon-Mom, W., Aartsma-Rus, A., and on behalf of the N = 1 Collaborative, U. (2024). Practical Recommendations for the Selection of Patients for Individualized Splice-Switching ASO-Based Treatments. Hum. Mutat. 2024, e9920230. doi: 10.1155/2024/9920230

Zerbino, D. R., Achuthan, P., Akanni, W., Amode, M. R., Barrell, D., Bhai, J., et al. (2018). Ensembl 2018. Nucleic Acids Res. 46, D754–D761. doi: 10.1093/nar/gkx1098

Zhao, S., Macakova, K., Sinson, J. C., Dai, H., Rosenfeld, J., Zapata, G. E., et al. (2025). Clinical validation of RNA sequencing for Mendelian disorder diagnostics. Am. J. Hum. Genet. 112, 779–792. doi: 10.1016/j.ajhg.2025.02.006

Zurek, B., Ellwanger, K., Vissers, L. E. L. M., Schüle, R., Synofzik, M., Töpf, A., et al. (2021). Solve-RD: systematic pan-European data sharing and collaborative analysis to solve rare diseases. Eur. J. Hum. Genet. EJHG 29, 1325–1331. doi: 10.1038/s41431-021-00859-0

