## Supplementary material for "Standardized transcriptome analysis improves rare disease diagnosis in the pan-European Solve-RD consortium": Sup Figures

### Supplementary Figures

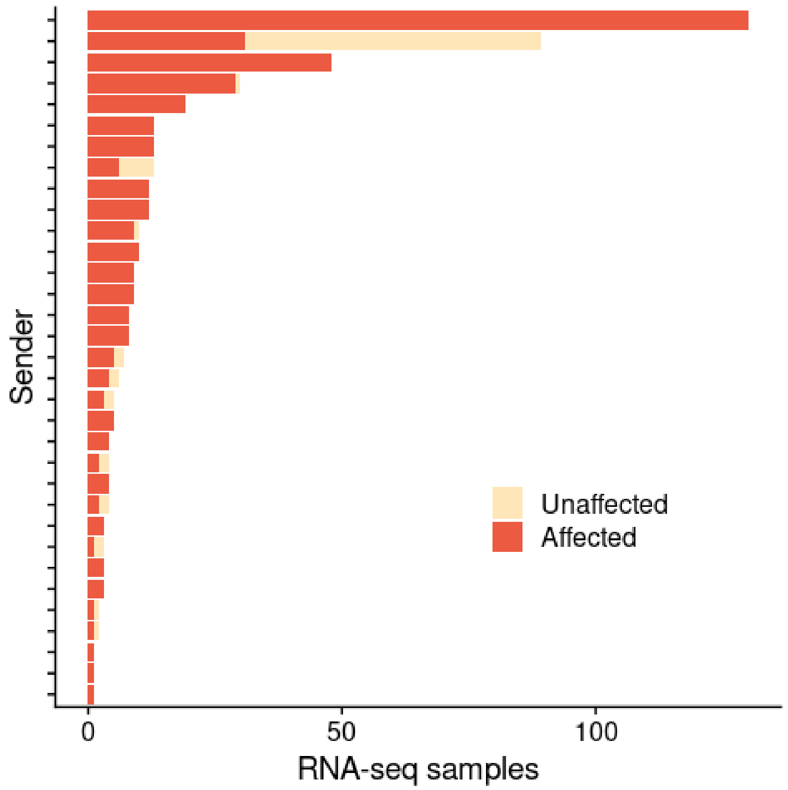


**Fig. S1. RNA-seq samples by sender.** Number of RNA-seq samples sent for analysis by each of the 33 different sender labs within Solve-RD. Unaffected samples come from family members.

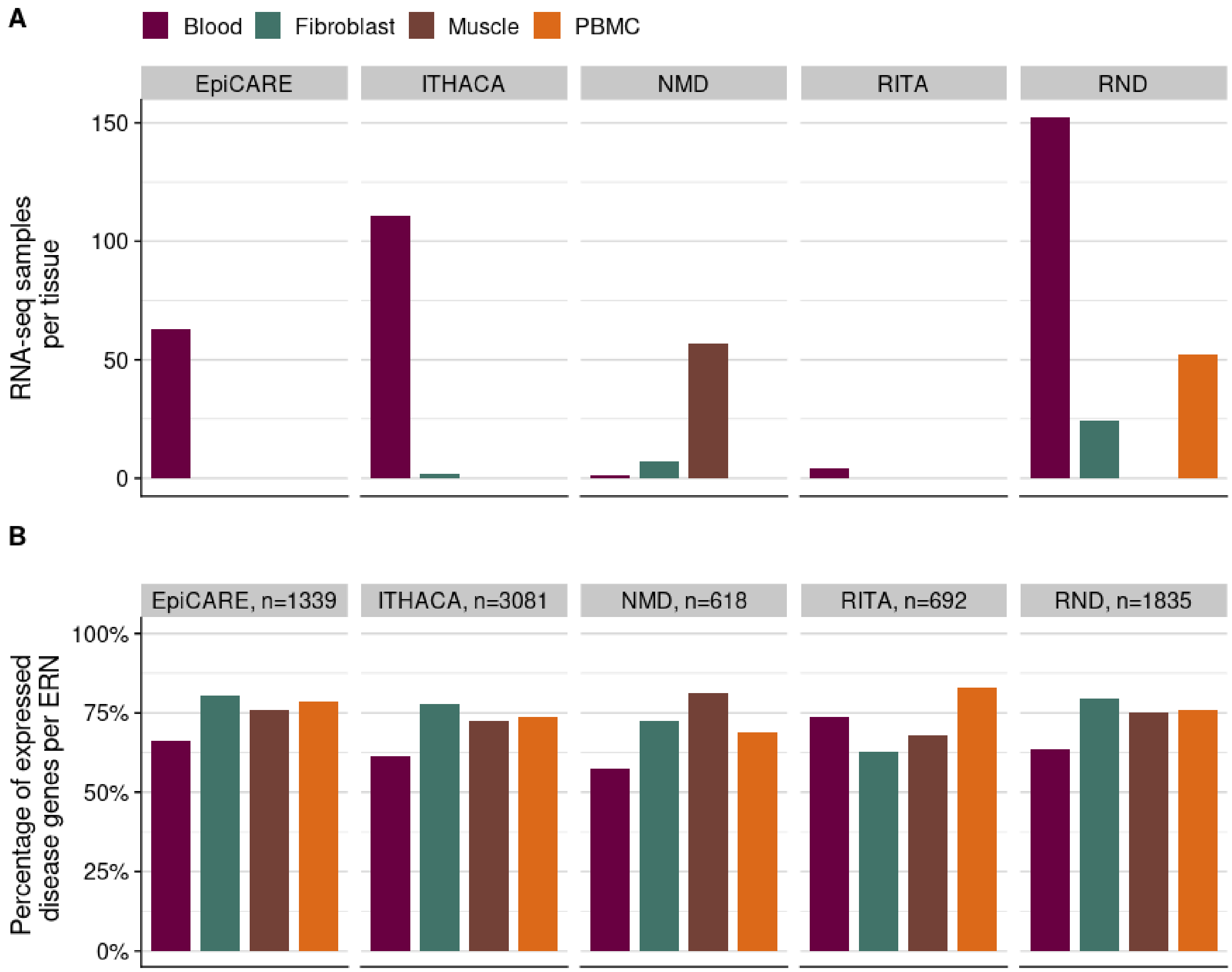


**Fig. S2. RNA-seq samples by tissue. A)** Number of RNA-seq samples sent by each ERN stratified by probed tissue. **B)** Percentage of expressed disease genes defined by each ERN stratified by tissue. n: denotes the number of disease genes.

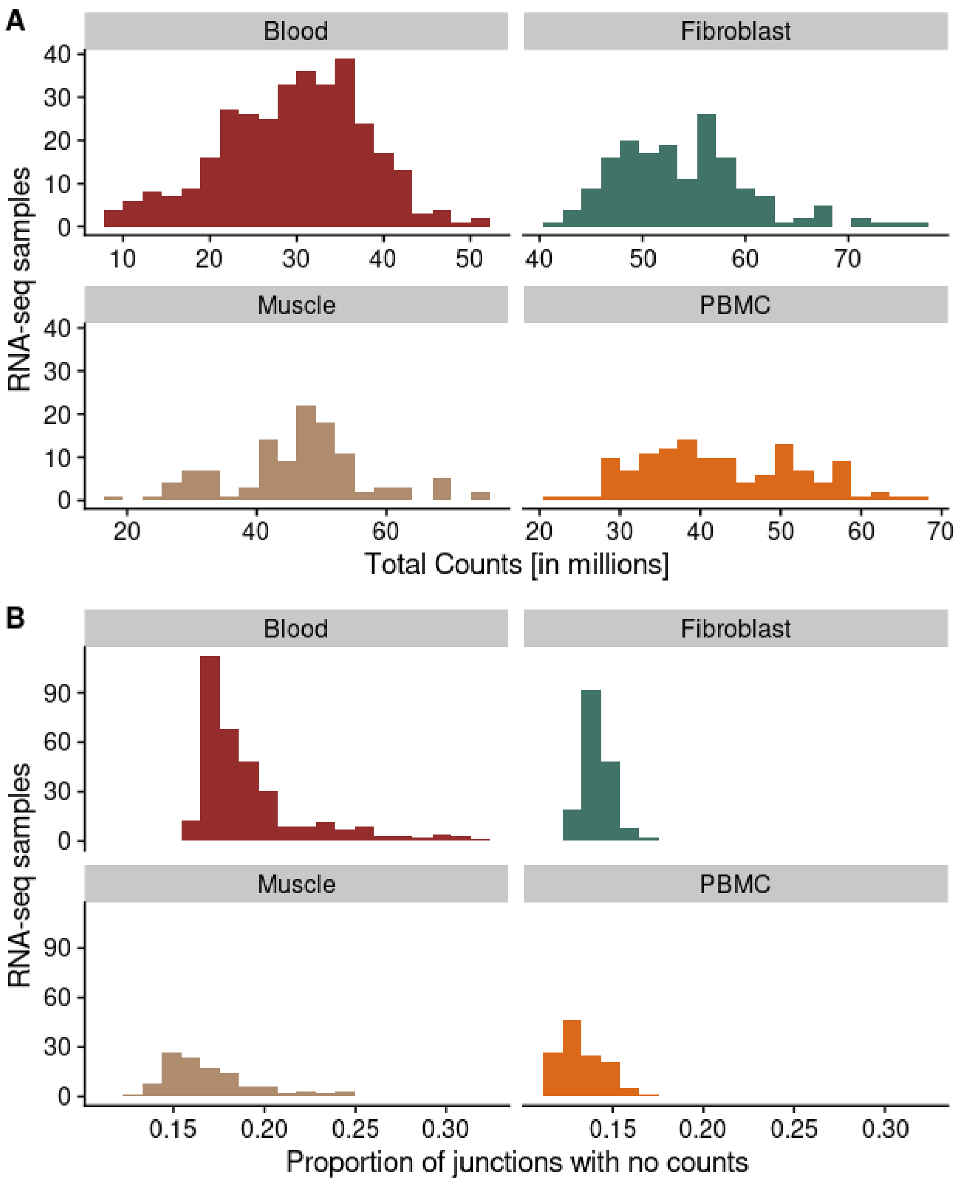


**Fig. S3. Read counts. A)** Total mapped counts [in millions] for each sample split by tissue. **B)** Proportion of junctions with no counts per tissue (junctions with no counts / total junctions that passed the filter).

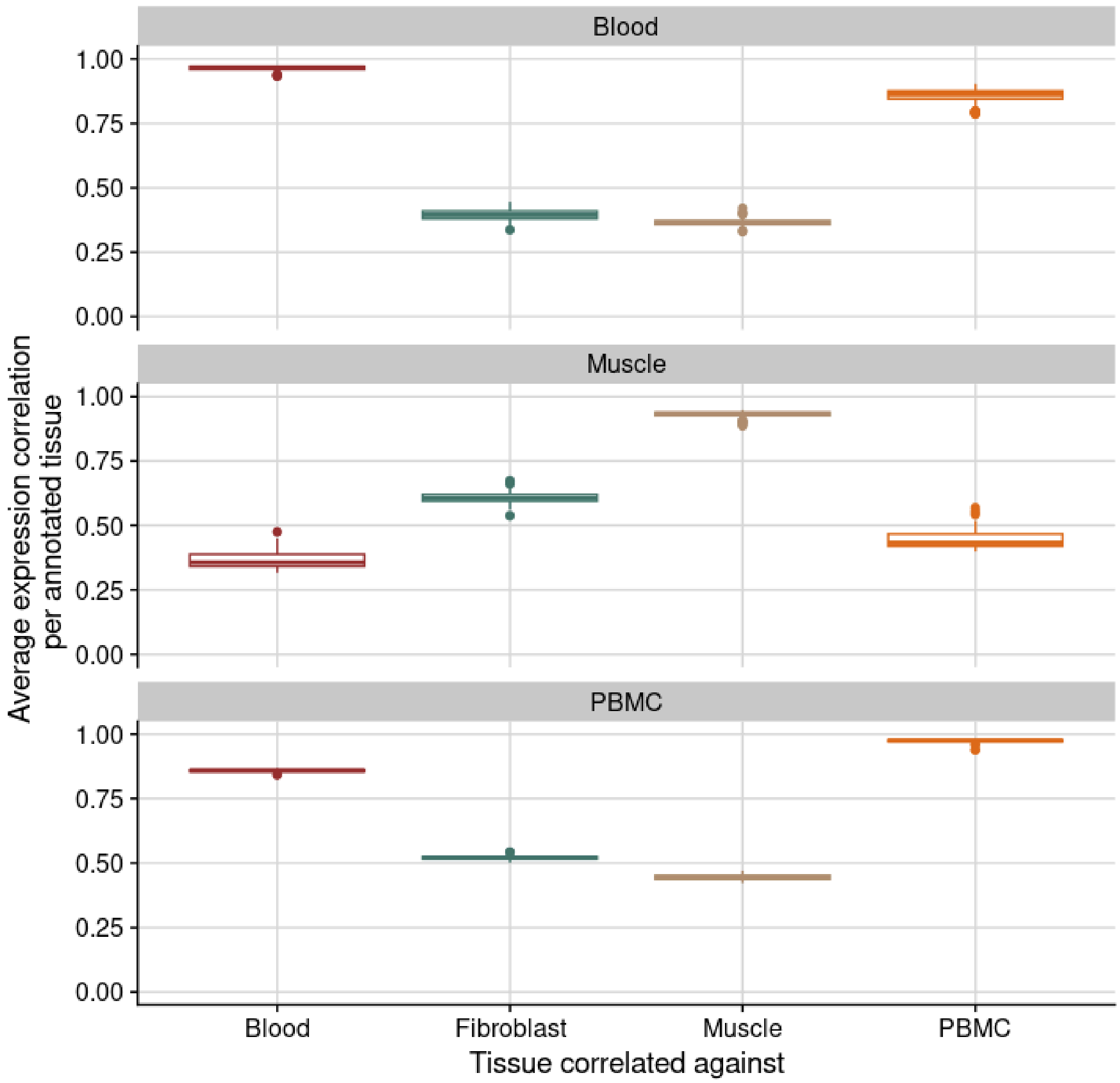


**Fig. S4. Sample correlation across tissues.** Average gene expression correlation (in FPKM) of all samples annotated as either blood, muscle, or PBMC against all the Solve-RD samples, by tissue.

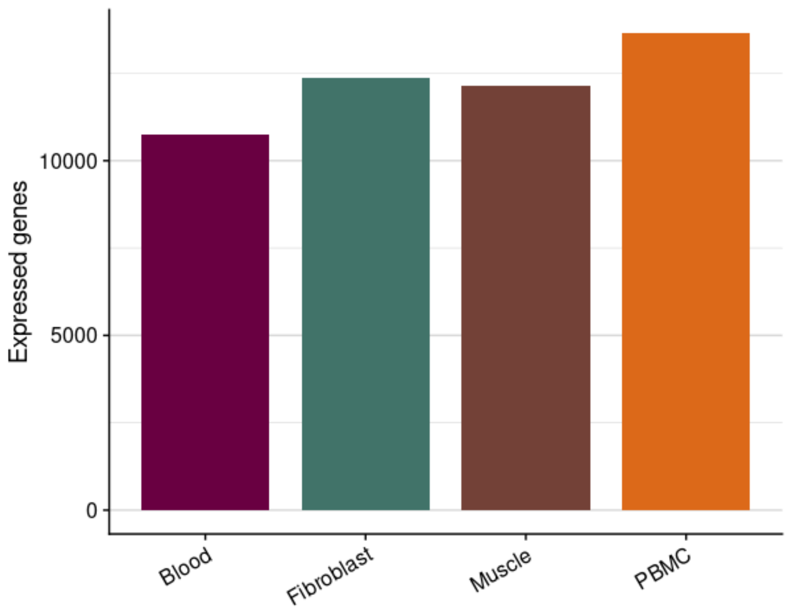


**Fig. S5. Number of expressed genes per tissue.**

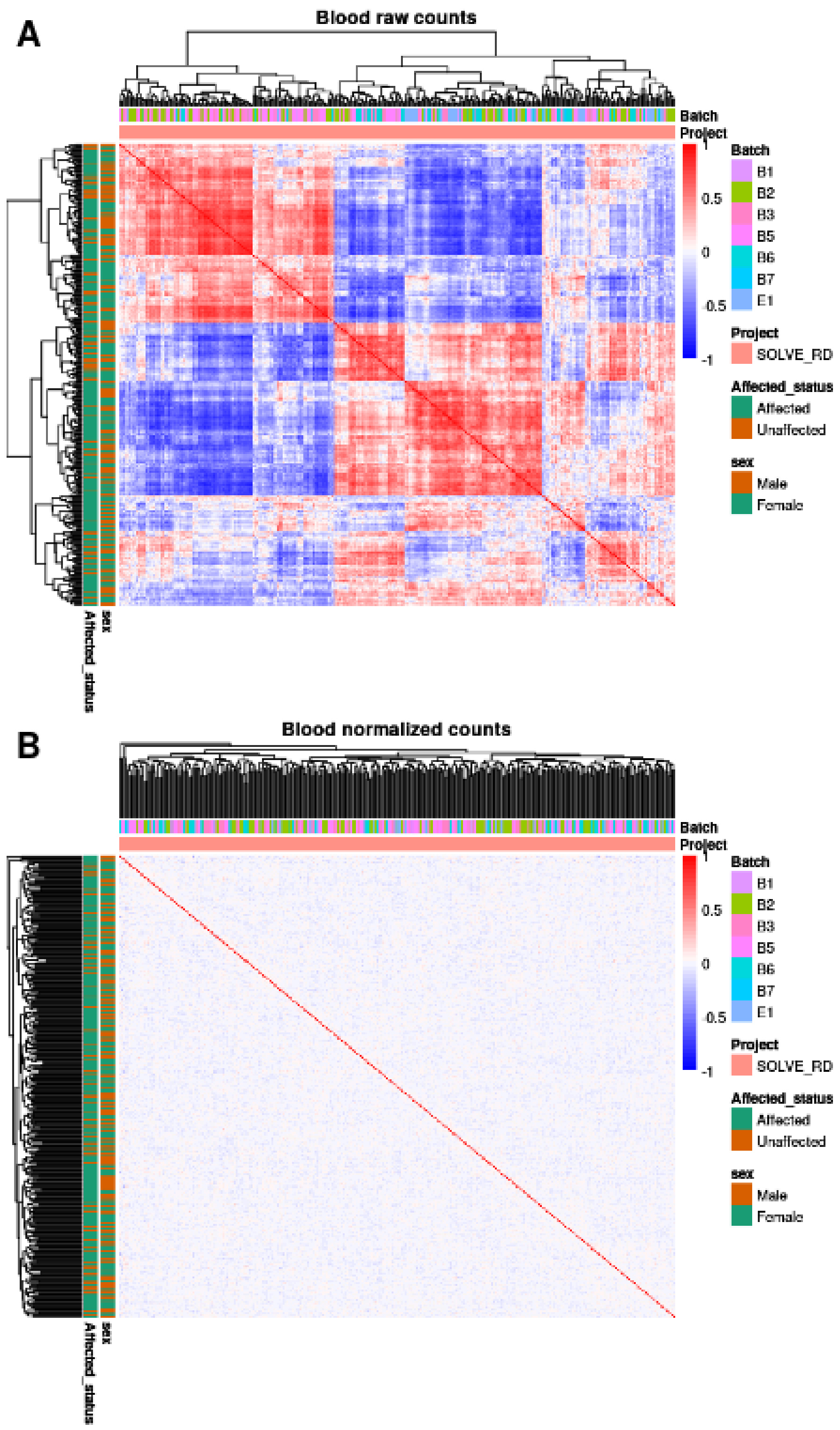


**Fig. S6.** Heatmap before and after correction for the blood cohort.

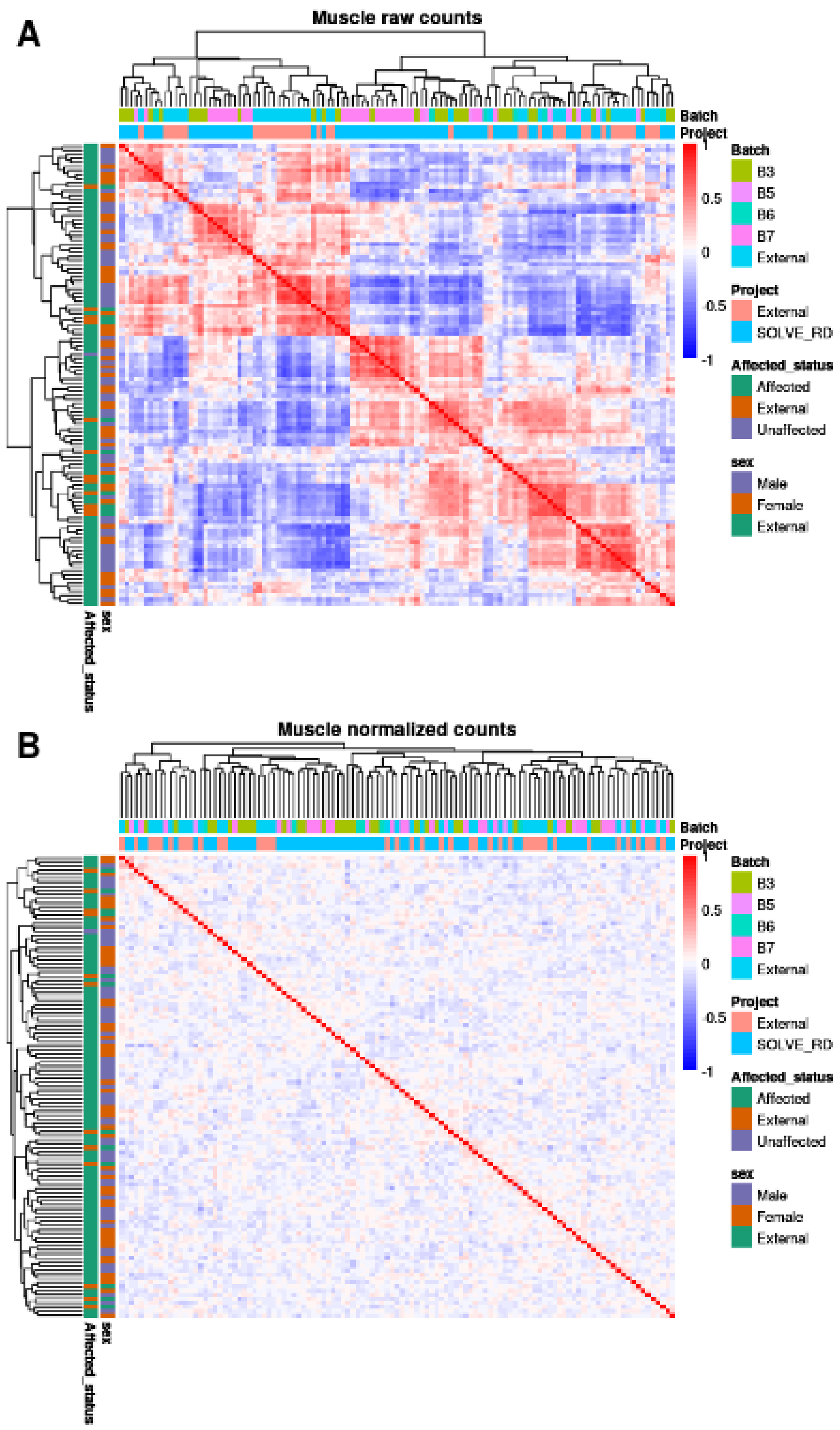


**Fig. S7.** Heatmap before and after correction for the muscle cohort.

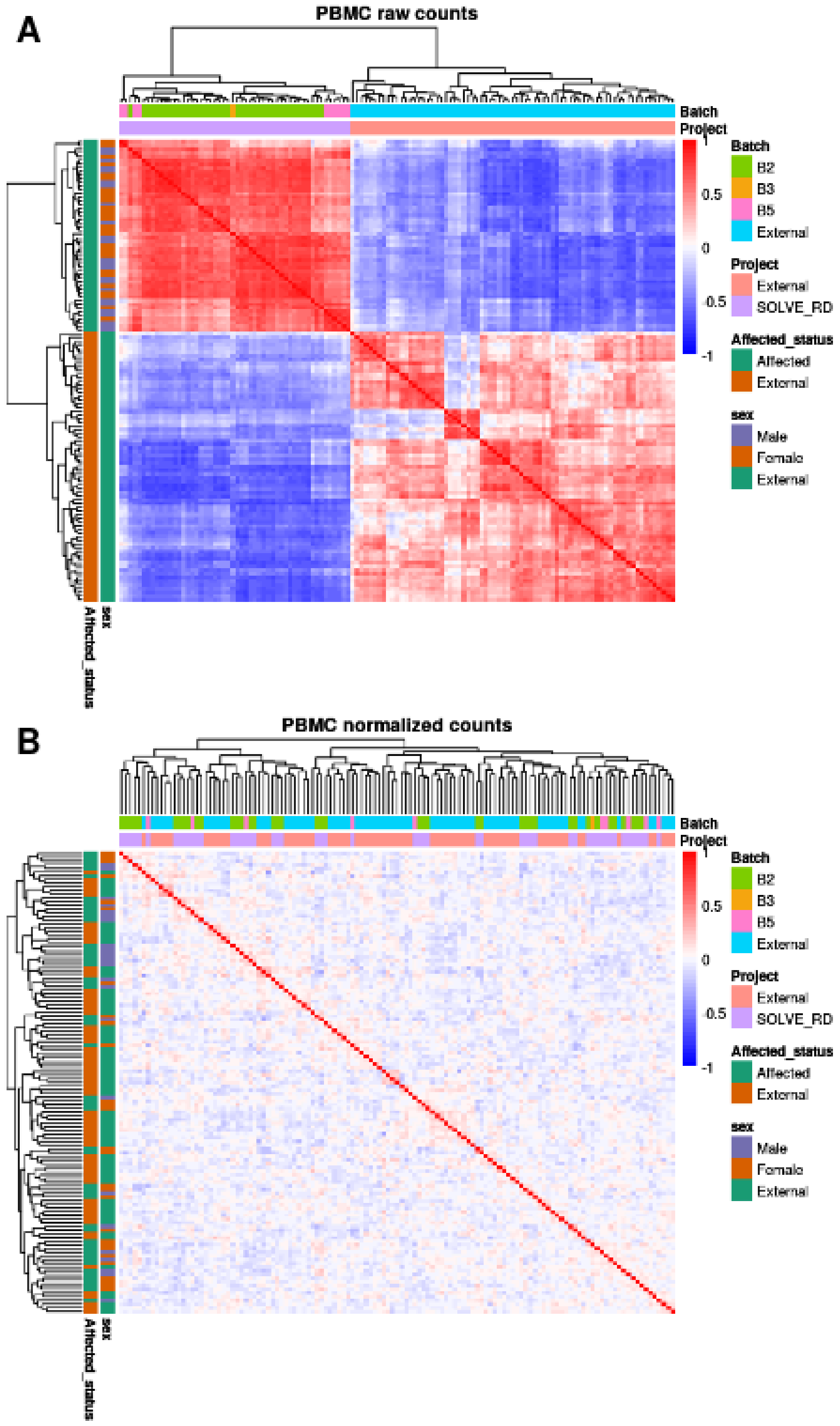


**Fig. S8.** Heatmap before and after correction for the PBMC cohort.

A

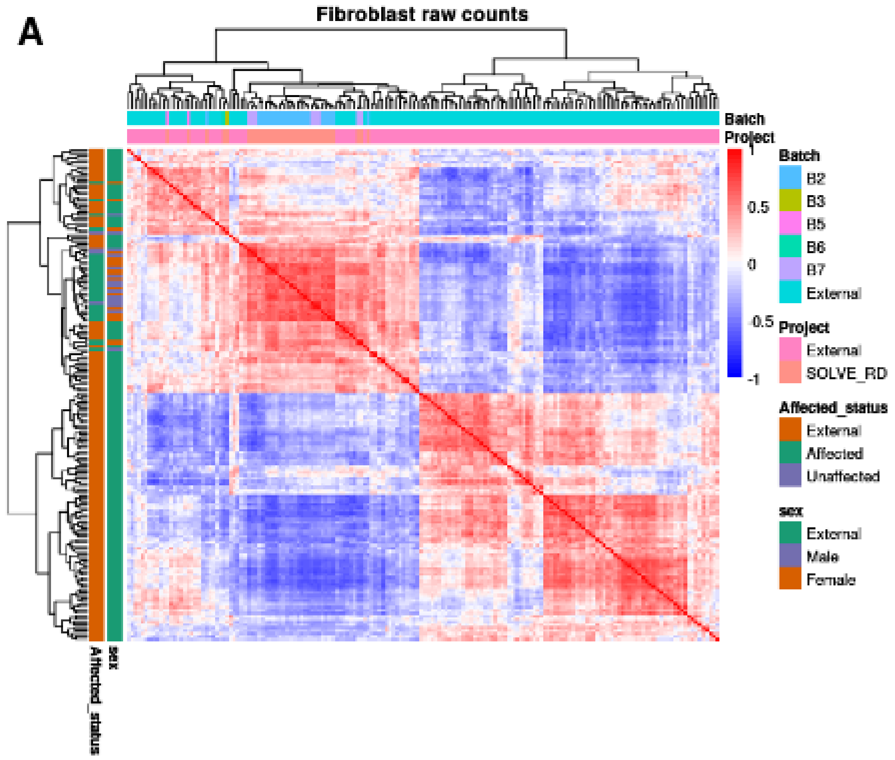


B

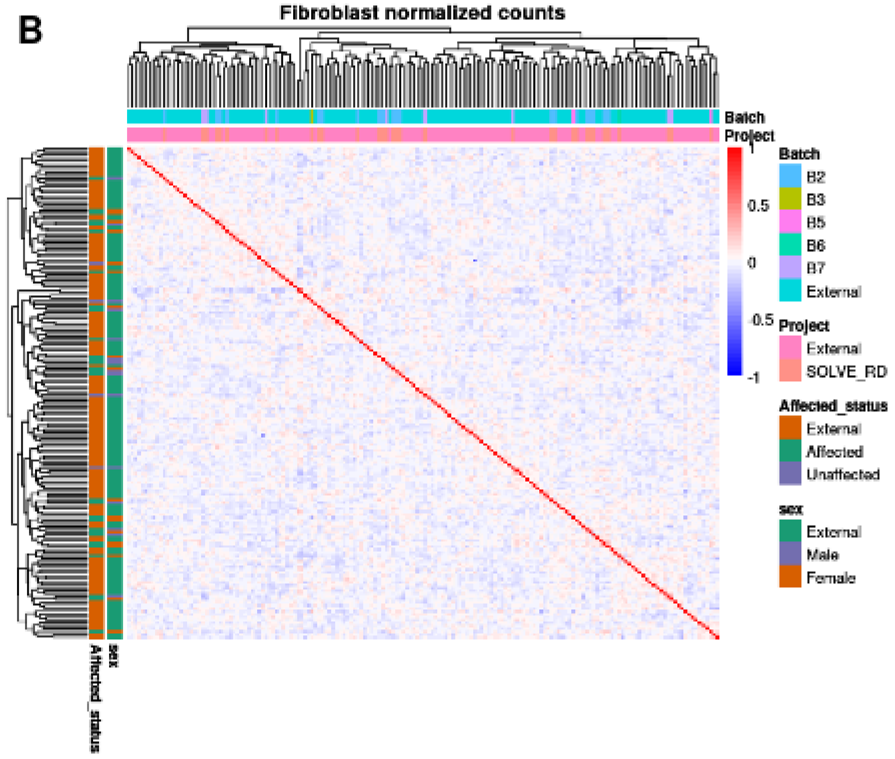


**Fig. S9.** Heatmap before and after correction for the fibroblast cohort.

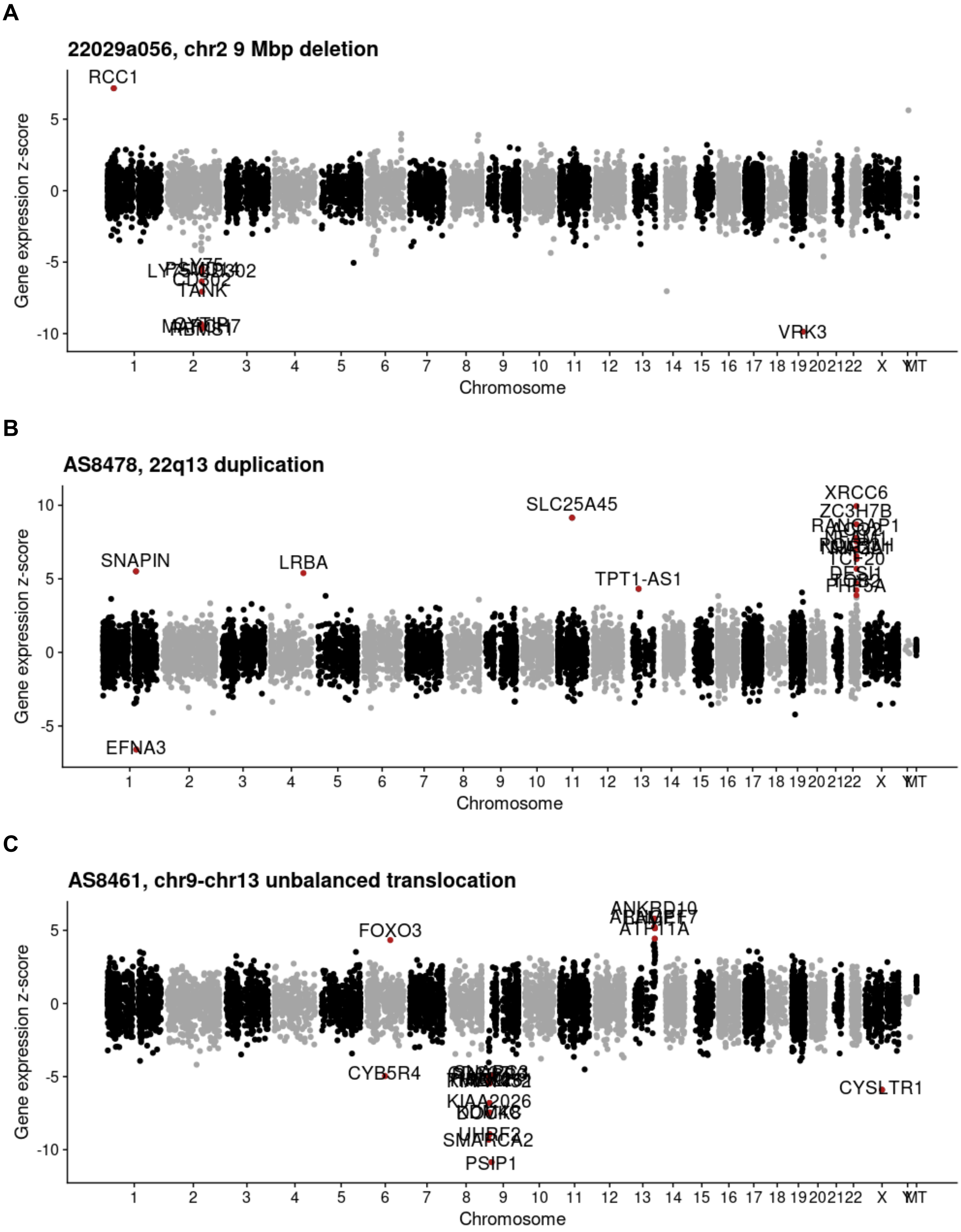
A
22029a056, chr2 9 Mbp deletion
RCC1
VRK3
TANK
B
AS8478, 22q13 duplication
SNAPIN
LRBA
SLC25A45
TPT1-AS1
XRCC6
ZC3H7B
EFNA3
C
AS8461, chr9-chr13 unbalanced translocation
FOXO3
ANKRD10
ATP11A
CYB5R4
KIAA2026
SMARCA2
PSIP1
CYSLTR1
Gene expression z-score
Chromosome

**Fig S10. Gene expression Manhattan plots.** Gene expression z-score against the genomic position of all expressed genes of 3 cases with disease-causal structural variants. Outlier genes in red.

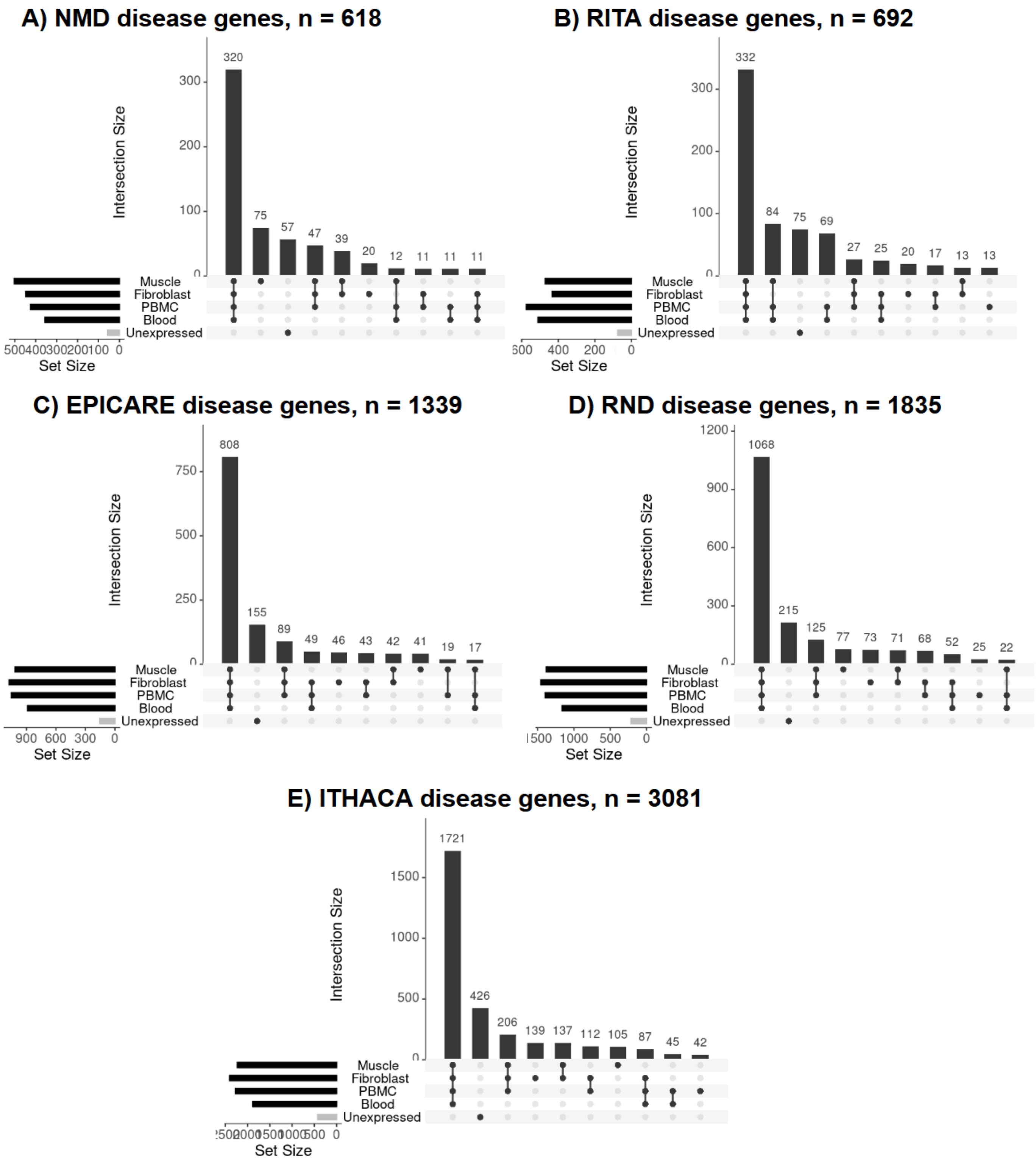


**Fig SX. Expression of ERN genes.** Upset plots showing the number of ERN-specific disease genes expressed in each tissue and in each tissue combination and also the number of unexpressed ERN genes.
